# Pre-existing autoimmunity is associated with increased severity of COVID-19: A retrospective cohort study using data from the National COVID Cohort Collaborative (N3C)

**DOI:** 10.1101/2023.02.02.23285353

**Authors:** Arjun S. Yadaw, Behdad Afzali, Nathan Hotaling, Hythem Sidky, Emily R Pfaff, David K. Sahner, Ewy A. Mathé, the N3C consortium

## Abstract

**Importance:** Identifying individuals with a higher risk of developing severe COVID-19 outcomes will inform targeted or more intensive clinical monitoring and management.

**Objective:** To examine, using data from the National COVID Cohort Collaborative (N3C), whether patients with pre-existing autoimmune disease (AID) diagnosis and/or immunosuppressant (IS) exposure are at a higher risk of developing severe COVID-19 outcomes.

**Design, setting and participants:** A retrospective cohort of 2,453,799 individuals diagnosed with COVID-19 between January 1^st^, 2020, and June 30^th^, 2022, was created from the N3C data enclave, which comprises data of 15,231,849 patients from 75 USA data partners. Patients were stratified as those with/without a pre-existing diagnosis of AID and/or those with/without exposure to IS prior to COVID-19.

**Main outcomes and measures:** Two outcomes of COVID-19 severity, derived from the World Health Organization severity score, were defined, namely life-threatening disease and hospitalization. Odds ratios (ORs) with 95% confidence intervals (CIs) were calculated using logistic regression models with and without adjustment for demographics (age, BMI, gender, race, ethnicity, smoking status), and comorbidities (cardiovascular disease, dementia, pulmonary disease, liver disease, type 2 diabetes mellitus, kidney disease, cancer, and HIV infection).

**Results:** In total, 2,453,799 (16.11% of the N3C cohort) adults (age> 18 years) were diagnosed with COVID-19, of which 191,520 (7.81%) had a prior AID diagnosis, and 278,095 (11.33%) had a prior IS exposure. Logistic regression models adjusted for demographic factors and comorbidities demonstrated that individuals with a prior AID (OR = 1.13, 95% CI 1.09 - 1.17; *p*=2.43E-13), prior exposure to IS (OR= 1.27, 95% CI 1.24 - 1.30; *p*=3.66E-74), or both (OR= 1.35, 95% CI 1.29 - 1.40; *p*=7.50E-49) were more likely to have a life-threatening COVID-19 disease. These results were confirmed after adjusting for exposure to antivirals and vaccination in a cohort subset with COVID-19 diagnosis dates after December 2021 (AID OR = 1.18, 95% CI 1.02 - 1.36; *p*=2.46E-02; IS OR= 1.60, 95% CI 1.41 - 1.80; *p*=5.11E-14; AID+IS OR= 1.93, 95% CI 1.62 - 2.30; *p*=1.68E-13). These results were consistent when evaluating hospitalization as the outcome and also when stratifying by race and sex. Finally, a sensitivity analysis evaluating specific IS revealed that TNF inhibitors were protective against life-threatening disease (OR = 0.80, 95% CI 0.66-0.96; *p*=1.66E-2) and hospitalization (OR = 0.80, 95% CI 0.73 - 0.89; *p*=1.06E-05).

**Conclusions and Relevance:** Patients with pre-existing AID, exposure to IS, or both are more likely to have a life-threatening disease or hospitalization. These patients may thus require tailored monitoring and preventative measures to minimize negative consequences of COVID-19.

## Introduction

The coronavirus disease of 2019 (COVID-19) pandemic has affected more than 664 million individuals and caused more than 6.7 million deaths worldwide, as of January 10^th^, 2023^1^. Most people who develop COVID-19 fully recover, but current evidence suggests approximately 10-20% of people experience a variety of mid- and long-term effects after they recover from their initial illness. These mid- and long-term effects are collectively known as post-acute sequelae of COVID-19 (PASC) or “long COVID”^2^. Thus, COVID-19 has had a major impact on human morbidity and mortality world-wide and represents a significant public health burden necessitating judicious allocation of healthcare resources. Indeed, the public health burden and the sheer magnitude of the pandemic underlie the importance of identifying patients at elevated risk of developing severe disease to inform targeted clinical monitoring and management. CDC guidelines provide a list of medical conditions, including, but not limited to, cancer, chronic kidney/liver/lung diseases and diabetes, that increase the risk of worse outcomes from COVID-19^3^. With the notable exception of type I diabetes, autoimmune diseases (AID) are excluded from this list. This is counter-intuitive, since these are common diseases (24 million people suffer from AID in the United States alone^4^), are typically life-long and incurable and are often treated with immunosuppressive drugs (IS), which could theoretically modify immunological responses to the SARS-CoV-2 virus. Therefore, it is important to directly evaluate, on a population-scale, the impact of AID and IS on severity outcomes of COVID-19. Such an evaluation would help inform healthcare guidelines and raise awareness for patients with AID so they can appropriately protect themselves from severe outcomes of COVID-19.

To date, there is mixed evidence regarding the association between AID and severity of outcomes from COVID-19. For example, SARS-CoV-2-infected patients with rheumatic and musculoskeletal diseases were reported to have a higher risk of developing COVID-19, be at a higher risk of hospitalization and severe (hospitalization or death) COVID-19, including requiring ICU admission and mechanical ventilation^5^. We also note that a recent study observed a higher risk of respiratory failure among patients with rheumatic disease with COVID-19^6–8^. In contrast, another report, a retrospective study of patients with AID hospitalized with COVID-19, did not show increased risk of an ICU admission, intubation, or death^9^. Another meta-analysis of observational and case-control studies, constrained to limited demographics (age, gender) and marked by considerable heterogeneity across studies, reported a high prevalence of COVID-19 in patients with AID, yet similar hospitalization and mortality rates of these patients compared to those without AID^10^. These studies are all limited by relatively small sample size, evaluating a limited number of AID, inadequately sampling representative populations, and/or failing to adjust for key confounders and known risk factors. Thus, whether AID are significant risk factors for worse outcomes from COVID-19 in larger cohorts that include a broad demographic and across the gamut of AID remains unknown.

An additional important confounder is whether immunosuppressive drugs also contribute to adverse outcomes from COVID-19. A recent study of 90 solid organ transplantation patients who were exposed to chronic immunosuppression and later diagnosed with COVID-19 had overall more severe disease^11^. Further, patients with cancer and solid-organ transplantation, and treated with IS for those conditions, may be at higher risk of severe COVID-19 outcomes, although patients with other AID may not be^12^, suggesting that immunosuppression may be just as relevant to COVID-19 outcomes as the underlying diseases. A recent study concluded that individuals on long term IS have worse outcomes when hospitalized with COVID-19 compared to those not on these medications^13^. Further, a meta-analysis of observational and case-control studies demonstrated that the use of glucocorticoids largely contributed to the association between AID and higher risk of COVID-19 infection. The same study demonstrated that exposure to glucocorticoids, conventional synthetic disease-modifying antirheumatic drugs (csDMARDs), or the combination of biologic or targeted synthetic DMARDs (b/tsDMARDS) and csDMARDS was associated with severe COVID-19 while b/tsDMARDs monotherapy (e.g. anti-TNF monotherapy) was associated with less severe COVID-19^10^. These findings imply that some forms of immunosuppressive therapies for AID could be protective against COVID-19, or less deleterious. Indeed, some clinical data suggest that prior treatment with TNF inhibitors may protect patients with psoriasis, at least compared with other forms of therapy^14^. Similarly, treatment with IL-1, IL-6, and JAK inhibitors *per se* are beneficial in patients with more severe COVID-19, and emerging data from the Accelerating COVID-19 Therapeutic Interventions and Vaccines study (ACTIV-1)^15^ also suggest anti-TNF therapy and abatacept may be beneficial in this context. Thus, a large-scale evaluation of IS in the context of AID to differentiate those that are protective from those that are harmful could help refine healthcare guidelines for patients using these medications.

To definitively establish whether individuals with AID or those treated with IS experience worse severity outcomes from COVID-19, we leveraged data from the N3C enclave, which harmonizes and holds electronic health records (EHRs) from 75 health systems with 15,231,849 million individuals’ data throughout the US, of which 5,858,748 have had COVID-19 ^16,17^. This represents the largest retrospective US cohort of SARS-CoV-2 patients. We hypothesized that patients with more severe COVID-19 outcomes (e.g., manifesting life-threatening disease or hospitalization) are more likely to have a prior diagnosis of an AID or have exposure to IS. We analyzed data made available between January 1^st^, 2020 and June 30^th^, 2022 and evaluated severity outcomes in patients with and without AID and/or IS prior to COVID-19, adjusting for key demographics and comorbidities. For a subset of this cohort (December 23rd, 2021 to June 30th, 2022), we also were able to adjust for vaccination status and antiviral treatment. Our analyses revealed that individuals with pre-existing AID, exposure to IS, or both are more likely to have severe COVID-19 outcomes, suggesting that these individuals may require careful monitoring and enhanced preventative measures to minimize short and long-term consequences of COVID-19.

## Methods

### Cohort Definition

The N3C enclave^18^ release version V90 from LDS data (N=15,231,849 patients), including individuals entered at or before August 25^th^ 2022 was used to define our cohort. We selected a subset of 2,453,799 patients that had a laboratory confirmed positive COVID-19 diagnosis based on a positive SARS-CoV-2 polymerase chain reaction (PCR) or antigen (Ag) test between January 1^st^ 2020 and June 30^th^ 2022 inclusive (**Figure 1**).

**Figure 1:**
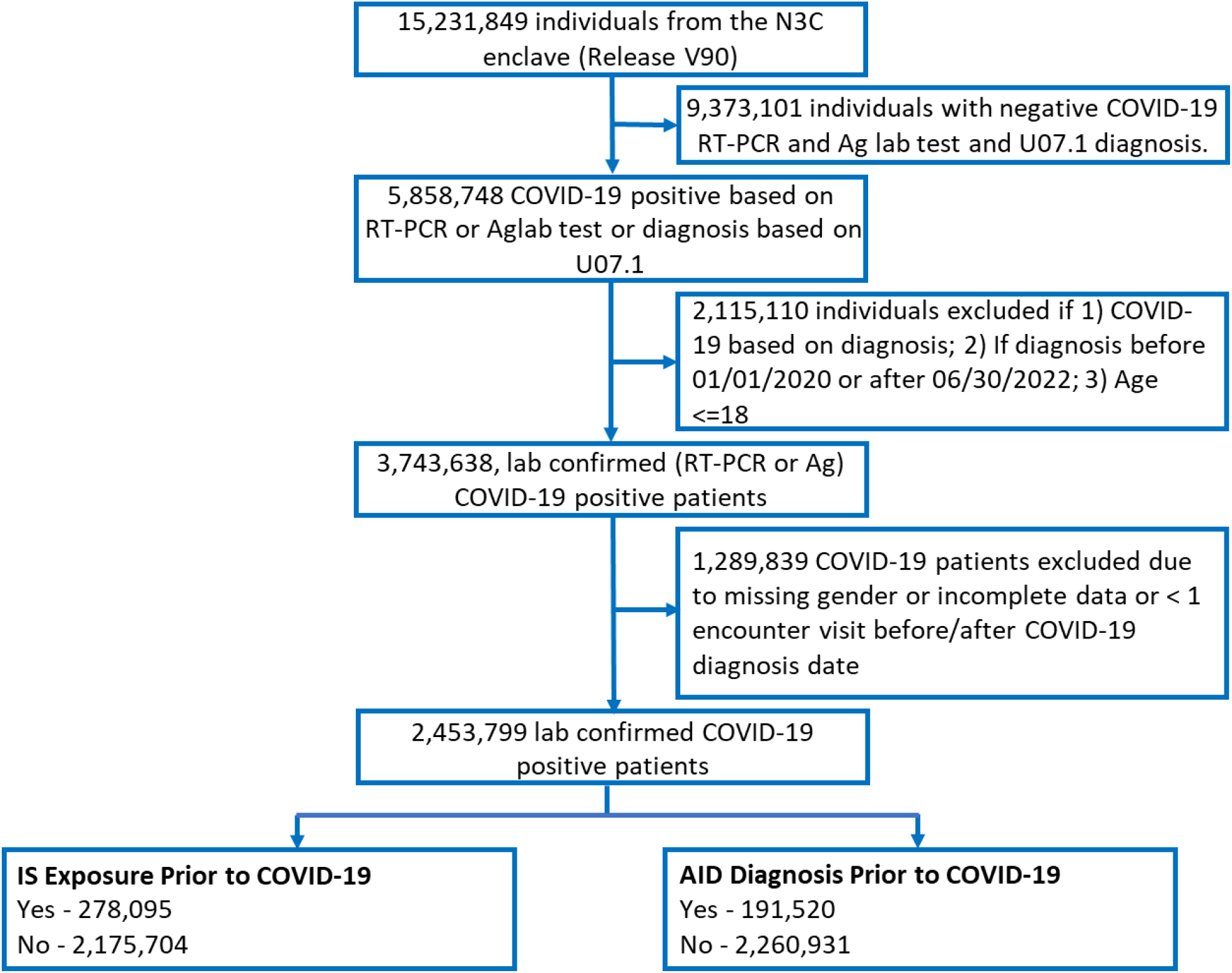
Workflow to define the AID cohort within N3C. A subset of 15,231,849 patients were identified in the N3C enclave release version V90. Of these, 2,453,799 were defined as COVID-19 positive between 2020-01-01 to 2022-06-30, as confirmed by a RT-PCR or Antigen test, and had non-missing values for ‘“age”‘ and “gender”. Patients were grouped based on whether or not they were diagnosed with an AID or exposed to IS prior to COVID-19. See Methods for further details on inclusion/exclusion criteria.

Each patient in the N3C enclave includes historical data from the same data partner dating from January 1, 2018 or later, thereby providing information on preexisting comorbidities, drug treatments, and other clinical information for many patients^13^. We excluded patients with missing age, missing gender, age <=18, with less than one encounter visit before or less than one encounter visit after COVID-19 diagnosis date, and patients from sites with data that did not meet quality check criteria.

Comorbidities of patients diagnosed with COVID-19 are reported in N3C as far back as 1^st^ January 2018 ^13,19^. Comorbidities were considered preexisting if their diagnosis date preceded that of COVID-19 diagnosis. Preexisting comorbidities considered include myocardial infarction (MI), congestive heart failure (CHF), peripheral vascular disease (PVD), stroke, dementia, pulmonary diseases, liver disease (mild and severe), type-2 diabetes mellitus, kidney disease, cancer (metastatic and non-metastatic), and human immunodeficiency virus (HIV) infection (eTable 1 in the **Supplement**). MI, CHF, PVD, and stroke were combined into a single cardiovascular disease (CVD) feature before inclusion as covariable in the adjusted analyses. Preexisting comorbidities were binarized (present or absent) for each condition and used as covariables in the adjusted statistical models.

### Severity Outcomes

COVID-19 severity outcomes defined in N3C are based on the Clinical Progression Scale (CPS) established by WHO^16,20^. Patients who are unaffected (WHO severity 0) were not considered in the analysis. For the remaining patients, severity of COVID-19 was classified as mild (WHO severity 1-3: outpatients with mild condition), mild_ED (WHO severity 3, outpatients with emergency department visit), moderate (WHO severity 4-6, hospitalized patients without invasive ventilation), severe (WHO severity 7-9, hospitalized patients with invasive ventilation or extracorporeal membrane oxygenation), mortality or hospice (WHO severity 10, hospital mortality or discharge to hospice). We further categorized the N3C severity outcomes as follows: 1) patients who experience, or not, life-threatening disease (Deceased/Severe vs. Moderate/Mild_ED/Mild); 2) patients hospitalized, or not (Dead/Severe/Moderate vs. Mild/Mild_ED).

### Definition of AID

A curated list of 106 AID based on two previously published lists^21,22^ was used to identify COVID-19 patients with or without AID within N3C (eFigure1A and eTable 2 in the **Supplement**). Each AID was mapped onto the OMOP (Observational Medical Outcomes Partnership) ontology as part of the N3C data harmonization procedure. Mapped AID were then grouped into a broader concept set manually with the help of a domain clinician expert. Patients with an AID diagnosis date prior to the COVID-19 diagnosis date were considered as having a preexisting AID. The first AID diagnosis date recorded was used for patients with multiple AID diagnosis dates.

### Definition of IS exposure

The following 15 drug classes, representing 303 different drugs^13^ (eFigure1B in the **Supplement**) were considered for defining COVID-19 patients with a prior IS exposure: Anthracyclines, Checkpoint Inhibitors, Cyclophosphamide, Protein Kinase Inhibitors, Rituximab, Monoclonal other, other antineoplastic agents (mAbs, Cancer Drugs L01 Other Cancer Therapies defined using WHO Anatomical Therapeutic Chemistry Class L01 products that were not anthracyclines, checkpoint inhibitors, cyclophosphamide, or protein kinase inhibitors), Azathioprine, Calcineurin Inhibitors, interleukin Inhibitors, JAK Inhibitors, Mycophenolate, TNF Inhibitors, Other selective immunosuppressants (Other Selective Immunosuppressants, L04 targeted cancer therapies defined using monoclonal antibody products in WHO Anatomical Therapeutic Chemistry Class L04 products that were not interleukin, tumor necrosis factor alpha or janus kinase inhibitors), Glucocorticoids. Patients diagnosed with COVID-19 were classified as having been exposed to IS if they were users of one or more of these medications at least 14 days prior to the COVID-19 diagnosis date^13^, and either continued with the medication during the COVID-19 visit or stopped on or after the date of diagnosis. Patients diagnosed with COVID-19 not satisfying these criteria were considered to not have been exposed to IS. “Of note, the computable phenotypes used in this manuscript differ from what has been previously published from this database as it was developed by the authors rather than the ISC domain team.” A subanalysis of AID patients with and without prior exposure to TNF inhibitors was performed, where TNF inhibitor exposure is defined as in the previous paragraph. For this analysis, patients with exposure to TNF inhibitors included those exposed to Etanercept, Infliximab, Afelimomab, Adalimumab, certolizumab pegol, Golimumab, or Opinercept.

### Definition of vaccination status and anti-viral usage

We selected a subcohort of patients diagnosed with COVID-19 between December 23rd 2021 and June 30th 2022 so that our models could be adjusted for vaccination status and exposure to antivirals. This time frame was selected because oral antiviral therapy became available through the FDA Emergency Use Authorization mechanism in late December 2021. In addition, December 2021 marked the onset of the Omicron phase of the pandemic, enabling restriction of the analysis to the single monoclonal antibody (bebtelovimab) with consistent activity against this dominant variant, at least through the end of the evaluation period, which preceded the emergence of subvariants of Omicron resistant to bebtelovimab. Only sites with vaccination rates matching CDC records for that site’s geographic region were included. Patients were considered vaccinated if they had at least one vaccination administered prior to COVID-19 diagnosis date. Patients exposed to antivirals were identified as those treated with at least one dose of any oral antiviral (Paxlovid (Nirmatrelvir/ritonavir), LAGEVRIO (molnupiravir)) or one monoclonal antibody (bebtelovimab) for COVID-19 between the first COVID-19 diagnosis date and up to 10 days before/thereafter. Of 2,453,799 COVID-19 patients, 248,743 were either vaccinated or treated with antivirals (eFigure1C in the **Supplement**).

### Statistical Analysis

Differences in clinical and demographic characteristics of study cohort patients with and without AID prior to COVID-19 diagnosis were analyzed using two-tailed Student’s t-test for continuous variables and Chi-square test for categorical variables. A series of logistic regression models, summarized in (eFigure2 in the **Supplement**), were then applied to test associations between prior exposure to AID, IS, or both with COVID-19 severity outcomes, independent of demographics, preexisting comorbidities, COVID-19 prevention/intervention (e.g. vaccination, antivirals), and TNF inhibition. Stratified models by race and preexisting cardiovascular diseases were also implemented.

Odds ratios derived from the coefficients of these logistic regression models were reported, along with 95% confidence intervals and p-values. Demographic variables include age, BMI, sex (male, female), race (Black or African American; White; Asian), ethnicity (Hispanic or Latino; not Hispanic or Latino), and smoking (non-smoker, current or former smoker). Preexisting comorbidities were also adjusted for as described above (see Cohort Definition). Logistic regression models were further stratified by race and gender. In a subset of the patients with available vaccination and anti-viral exposure data, the same base logistic regression models were used with the addition of vaccination and anti-viral exposure status as covariables.

All statistical modeling was conducted within the N3C enclave by using SQL, Python(3.6.7), statsmodels (version 0.12.2), Patsy (version 0.5.2), and scipy(1.6.2). Statistical significance was defined for p-values < 0.05 and 95% confidence intervals around the estimated odd ratios are reported. The baseline characteristic table 1 was created by using the tableone python package^23^.

**Table 1:** Characteristics of COVID-19 positive patients with and without autoimmune diseases (AID)/Immunosuppressants (IS) usage. All p-values are <0.001 for all continuous and categorical variables (cohorts with/without AID and cohort with/without IS).

|  | Overall N3C sample<br>(n = 2,453,799) | Patients without AID prior COVID-19<br>(n = 2,262,279) | Patients with AID prior to COVID-19<br>(n = 191,520) | Patients without IS prior to COVID-19<br>(n = 2,175,704) | Patients with IS prior to COVID-19<br>(n = 278,095) |
| --- | --- | --- | --- | --- | --- |
| **COVID-19 severity** |  |  |  |  |  |
| Life threatening, n(%) | 54932 (2.2) | 46335 (2.0) | 8597 (4.5) | 41212 (1.9) | 13720 (4.9) |
| Hospitalized, n(%) | 220353 (9.0) | 189519 (8.4) | 30834 (16.1) | 174587 (8.0) | 45766 (16.5) |
| **Demographic** |  |  |  |  |  |
| Age (years), mean (SD) | 47.4 (18.0) | 46.9 (18.0) | 53.8 (17.2) | 46.8 (18.0) | 52.3 (17.4) |
| BMI (kg/m<sup>2</sup>), mean (SD) | 32.3 (8.4) | 32.2 (8.3) | 32.9 (8.6) | 31.8 (8.2) | 33.9 (9.0) |
| Gender, n(%) |  |  |  |  |  |
| Female | 1454673 (59.3) | 1323247 (58.5) | 131426 (68.6) | 1281335 (58.9) | 173338 (62.3) |
| Male | 999126 (40.7) | 939032 (41.5) | 60094 (31.4) | 894369 (41.1) | 104757 (37.7) |
| Race, n(%) |  |  |  |  |  |
| Black or African American | 342511 (14.0) | 316416 (14.0) | 26095 (13.6) | 295376 (13.6) | 47135 (16.9) |
| White | 1790481 (73.0) | 1644325 (72.7) | 146156 (76.3) | 1590559 (73.1) | 199922 (71.9) |
| Asian | 61597 (2.5) | 57870 (2.6) | 3727 (1.9) | 56531 (2.6) | 5066 (1.8) |
| Native Hawaiian or Other Pacific Islander | 3940 (0.2) | 3691 (0.2) | 249 (0.1) | 3499 (0.2) | 441 (0.2) |
| Missing/Unknown/Other | 255270 (10.4) | 239977 (10.6) | 15293 (8.0) | 229739 (10.6) | 25531 (9.2) |
| Ethnicity, n(%) |  |  |  |  |  |
| Hispanic or Latino | 269354 (11.0) | 252829 (11.2) | 16525 (8.6) | 244580 (11.2) | 24774 (8.9) |
| Not Hispanic or Latino | 1966475 (80.1) | 1805569 (79.8) | 160906 (84.0) | 1739697 (80.0) | 226778 (81.5) |
| Missing/Unknown | 217970 (8.9) | 203881 (9.0) | 14089 (7.4) | 191427 (8.8) | 26543 (9.5) |
| Smoking status, n(%) |  |  |  |  |  |
| Non-smoker | 2173346 (88.6) | 2014857 (89.1) | 158489 (82.8) | 1959001 (90.0) | 214345 (77.1) |
| Current or former smoker | 280453 (11.4) | 247422 (10.9) | 33031 (17.2) | 216703 (10.0) | 63750 (22.9) |
| **Comorbidities** |  |  |  |  |  |
| Cardiovascular disease, n(%) | 296990 (12.1) | 237822 (10.5) | 54154 (28.3) | 216891 (10.0) | 80099 (28.8) |
| Dementia, n(%) | 32023 (1.3) | 26950 (1.2) | 5073 (2.6) | 25810 (1.2) | 6213 (2.2) |
| Chronic pulmonary disease, n(%) | 333222 (13.6) | 281063 (12.4) | 52159 (27.2) | 242727 (11.2) | 90495 (32.5) |
| Liver mild, n(%) | 113751 (4.6) | 91069 (4.0) | 22682 (11.8) | 79800 (3.7) | 33951 (12.2) |
| Liver severe, n(%) | 13463 (0.5) | 9817 (0.4) | 3646 (1.9) | 8378 (0.4) | 5085 (1.8) |
| Type 2 diabetes, n(%) | 308690 (12.6) | 255620 (11.3) | 53070 (27.7) | 242472 (11.1) | 66218 (23.8) |
| Kidney, n(%) | 138958 (5.7) | 108921 (4.8) | 30037 (15.7) | 97804 (4.5) | 41154 (14.8) |
| Cancer, n(%) | 137706 (5.6) | 115144 (5.1) | 22562 (11.8) | 95543 (4.4) | 42163 (15.2) |
| MetS, n(%) | 22737 (0.9) | 19051 (0.8) | 3686 (1.9) | 12304 (0.6) | 10433 (3.8) |
| HIV, n(%) | 10959 (0.4) | 9882 (0.4) | 1077 (0.6) | 8625 (0.4) | 2334 (0.8) |

Abbreviations: AID, autoimmune disease; IS, Immunosuppressants; BMI, body mass index ; MetS, Metastatic cancer; HIV, Human Immunodeficiency virus; Cardiovascular disease (Myocardial infarction or Congestive heart failure or Peripheral vascular disease or Stroke); Life-threatening (Death, ECMO or mechanical ventilation) vs (Moderate or Mild\_ED or Mild); Hospitalized: (Death, ECMO or mechanical ventilation or Moderate) vs (Mild\_ED or Mild).

## Results

### Cohort Description

We defined a large cohort within the N3C data enclave^18^ to evaluate the impact of prior AID diagnosis and exposure to IS on COVID-19 severity outcomes (**Figure 1** and eFigure1 in the **Supplement**). Using the N3C dataset released on Aug 25^th^, 2022, comprising a total of 15,231,849 individuals, 2,453,799 patients diagnosed with COVID-19 were identified, as indicated by a positive RT-PCR or antigen test between Jan 1^st^, 2020 and June 30^th^ 2022 inclusive. Among the 2,453,799 patients diagnosed with COVID-19, 220,353 (9%) were hospitalized and 54,932 (2.2%) had life-threatening disease. Patients were further categorized as those with a pre-existing AID (N=191,520 patients), those with exposure to IS (N=278,095 patients), and those with both AID and IS (N=56,813) prior to COVID-19 diagnosis (**Figure 1;** eFigure1A & eFigure1B in the **Supplement**).

Demographic characteristics and preexisting comorbidities (prior to COVID-19 diagnosis) of patients diagnosed with COVID-19 with and without preexisting AID diagnosis or exposure to IS are shown in **Table 1**. There were more female (N=131,426; 68.6%) and less male (N=60,094; 31.4%) patients with COVID-19 and prior AID compared to those without AID (1,323,247 females [58.5%] and 939,032 males [41.5%]). Patients with COVID-19 and AID were older than those without AID (mean(s.d.) 53.8(17.2) vs. 46.9(18.0) years (*P*< 0.001). Patients with prior AID also had significantly higher prevalence for all preexisting comorbidities, including type-2 diabetes, chronic pulmonary disease and liver disease (see **Table 1**).

The top 3 most abundant AID were rheumatoid arthritis (N=27,664), psoriasis (N=25,749), and Type I diabetes mellitus (N=24,443). The top 20 most abundant AID are shown in **Figure 2A**. Interestingly, the most prevalent conditions showed a larger proportion of patients with life-threatening disease or hospitalization (eFigure3A & eFigure3C in the **Supplement**). The top three most frequent IS drugs that patients were exposed to were glucocorticoids, calcineurin inhibitors, and other antineoplastic agents (mAbs, Cancer Drugs L01 Other Cancer Therapies defined using WHO Anatomical Therapeutic Chemistry Class L01 products that were not anthracyclines, checkpoint inhibitors, cyclophosphamide, or protein kinase inhibitors) (**Figure 2B**). Most patients with COVID-19 have a single preexisting AID diagnosis (159,770) and an exposure to a single IS (n=237,238 patients, representing 85.31% of patients with IS exposure) (**Figure 2C**). Lastly, we note that the most frequent IS drugs showed a larger proportion of patients with life-threatening conditions or hospitalization (eFigure3B & eFigure3D in the **Supplement**).

**Figure 2:**
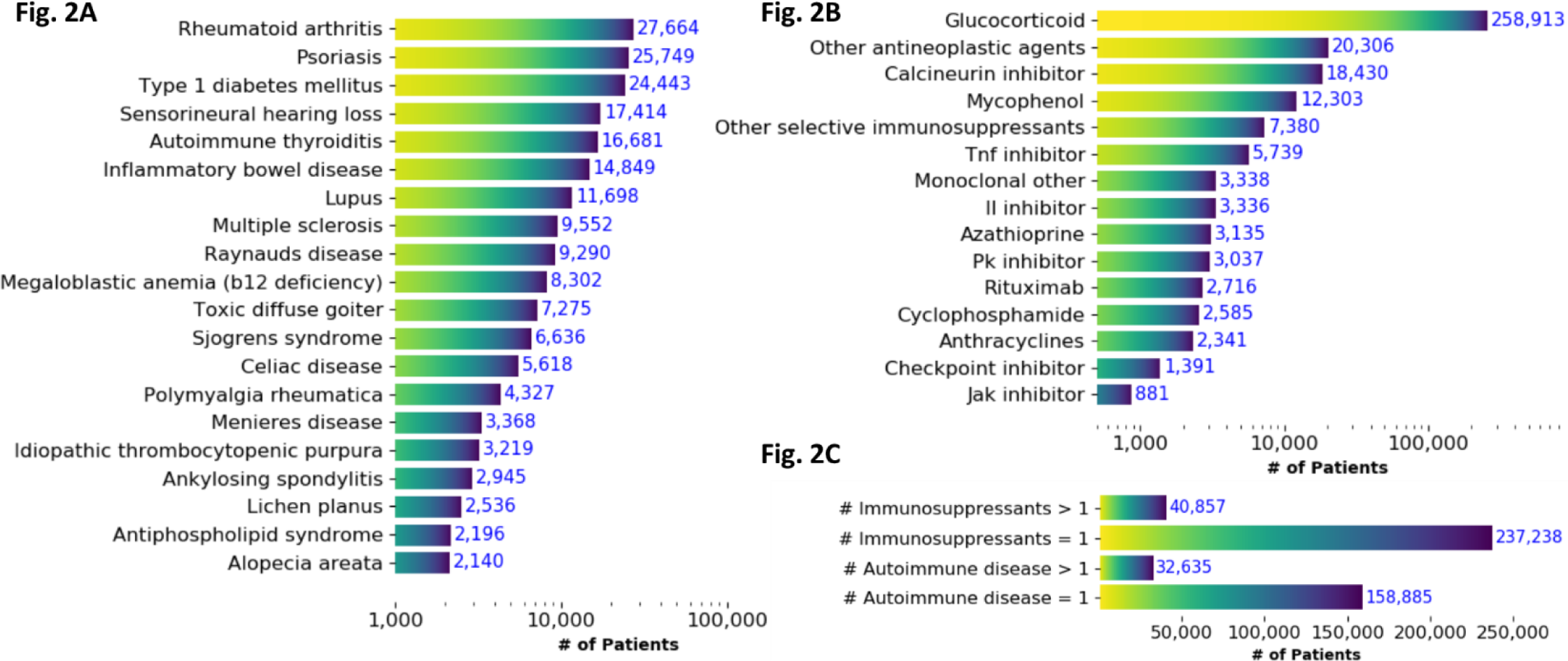
Description of Cohort Exposures. Top 20 most abundant AID (A) and immunosuppressant exposures (B) prior to COVID-19 diagnosis of lab confirmed COVID-19 patients. “L04 other” include “Selective immunosuppressants” that were not IL inhibitors, JK inhibitors, TNF alpha inhibitors or monoclonal antibodies. “L01 other” includes “Other cancer therapies” not anthracyclines, checkpoint inhibitors, cyclophosphamide, or protein kinase inhibitors. (C) Number of patients with single/multiple AIDs and patients single/multiple immunosuppressants exposure

### Association between prior exposure to AID, IS, or both (AID-IS) with COVID-19 severity outcomes

Two binary clinically relevant COVID-19 severity outcomes were defined: 1. presence or absence of life-threatening disease and 2. hospitalization with COVID-19 (**see Methods**). We tested whether the two outcomes were more likely in the setting of a preexisting diagnosis of AID or exposure to IS or both, using univariate and multivariate models adjusted for demographics (eTable 3 in the **Supplement**). In our final multivariate regression analyses (**Table 2**), the model was adjusted for basic demographic characteristics (age, BMI, sex, race, ethnicity, smoking) as well as known comorbidities that increase the risk of more severe COVID-19 infection (CVD, dementia, pulmonary disease, liver disease, cancer, Type-2 diabetes, kidney disease, cancer, and HIV infection). In both univariate and multivariate analysis, we found that patients with a prior diagnosis of AID, prior exposure to IS or both pre-existing AID *and* exposure to IS were more likely to have more severe outcomes than patients with neither AID nor IS (**Table 2**). When adjusting for demographic factors and comorbidities, patients were almost 21% more likely to be hospitalized if they had pre-existing AID (OR = 1.21, 95% CI 1.19 - 1.24, *P* =2.85E-97), 19% more likely if they had prior exposure to IS (OR= 1.19, 95% CI 1.17 - 1.21, *P* = 2.28E-120), or 31% more likely if they had both (OR= 1.31, 95% CI 1.28 - 1.34, *P* = 8.11E-109). Similarly, when adjusting for demographics and comorbidities, patients were 13% more likely to develop life-threatening COVID-19 if they had pre-existing AID (OR = 1.13, 95% CI 1.10 - 1.17, *P* = 2.43E-13), 27% more likely if they had prior exposure to IS (OR= 1.27, 95% CI 1.24 - 1.30, *P* = 3.66E-74), or 35% more likely if they had both (OR= 1.35, 95% CI 1.29 - 1.40, *P* = 7.50E-49) (**Table 2**).

**Table 2:** Multivariate logistic regression model of severity outcomes adjusted for demographics and comorbidities (January 1, 2020, to June 30, 2022, n = 2,453,799)

|  | Life-threatening condition;<br>(Yes: 54,932,<br>No: 2,398,867)<br>OR (95% CI) | P value | Hospitalized condition;<br>(Yes: 220,353,<br>No: 2,233,446)<br>OR (95% CI) | P value |
| --- | --- | --- | --- | --- |
| **AID/Immunosuppressants (IS) status** |  |  |  |  |
| AID only | 1.13 (1.09 - 1.17) | 2.43E-13 | 1.21 (1.19 - 1.24) | 2.85E-97 |
| IS only | 1.27 (1.24 - 1.30) | 3.66E-74 | 1.19 (1.17 - 1.21) | 2.28E-120 |
| AID + IS only | 1.35 (1.29 - 1.40) | 7.50E-49 | 1.31 (1.28 - 1.34) | 8.11E-109 |
| **Demographics** |  |  |  |  |
| **Age** | 2.84 (2.81 - 2.88) | <2.2E-308 | 1.94 (1.93 - 1.95) | <2.2E-308 |
| **BMI** | 1.08 (1.07 - 1.09) | 4.77E-72 | 1.10 (1.09 - 1.11) | <2.2E-308 |
| **Gender** |  |  |  |  |
| Female | 1 [Reference] |  | 1 [Reference] |  |
| Male | 1.51 (1.48 - 1.53) | <2.2E-308 | 1.29 (1.28 - 1.30) | <2.2E-308 |
| **Race** |  |  |  |  |
| White | 1 [Reference] |  | 1 [Reference] |  |
| Others/unknown | 1.20 (1.16 - 1.25) | 1.24E-24 | 1.18 (1.16 - 1.20) | 1.66E-74 |
| Black or African American | 1.37 (1.33 - 1.40) | 3.07E-131 | 1.77 (1.75 - 1.80) | <2.2E-308 |
| Race asian | 1.32 (1.24 - 1.40) | 2.68E-18 | 1.35 (1.31 - 1.40) | 2.38E-78 |
| **Ethnicity** |  |  |  |  |
| Not Hispanic or Latino | 1 [Reference] |  | 1 [Reference] |  |
| Hispanic or Latino | 1.18 (1.13 - 1.22) | 1.93E-18 | 1.39 (1.36 - 1.41) | 3.61E-304 |
| **Smoking status** |  |  |  |  |
| Nonsmoker | 1 [Reference] |  | 1 [Reference] |  |
| Current or Former | 1.37 (1.34 - 1.40) | 6.48E-154 | 1.61 (1.59 - 1.63) | <2.2E-308 |
| **Comorbidities** |  |  |  |  |
| Cardiovascular disease | 1.69 (1.66 - 1.73) | <2.2E-308 | 1.67 (1.65 - 1.69) | <2.2E-308 |
| Dementia | 2.22 (2.16 - 2.30) | <2.2E-308 | 1.99 (1.94 - 2.04) | <2.2E-308 |
| Chronic pulmonary disease | 1.23 (1.21 - 1.26) | 1.23E-81 | 1.29 (1.27 - 1.30) | <2.2E-308 |
| Liver mild | 1.34 (1.30 - 1.38) | 3.59E-78 | 1.25 (1.22 - 1.27) | 6.41E-124 |
| Liver severe | 3.05 (2.89 - 3.23) | <2.2E-308 | 2.33 (2.23 - 2.42) | <2.2E-308 |
| Kidney disease | 1.77 (1.73 - 1.81) | <2.2E-308 | 1.88 (1.86 - 1.91) | <2.2E-308 |
| Cancer | 1.45 (1.41 - 1.49) | 7.29E-173 | 1.24 (1.22 - 1.26) | 2.30E-137 |
| Metastatic cancer | 3.31 (3.17 - 3.46) | <2.2E-308 | 2.19 (2.12 - 2.26) | <2.2E-308 |
| Type 2 Diabetes Mellitus | 1.32 (1.29 - 1.35) | 1.19E-149 | 1.46 (1.44 - 1.48) | <2.2E-308 |
| HIV | 1.24 (1.12 - 1.38) | 5.48E-05 | 1.26 (1.19 - 1.33) | 2.16E-16 |

Demographic factors include age, BMI, gender, race, ethnicity, smoking status.
Comorbidities include cardiovascular disease (MI + CHF + PVD + stroke), dementia, pulmonary disease, liver disease (mild and severe), Type 2 diabetes, kidney disease, cancer (metastatic and non-metastatic), and HIV infection.

### Association between AID, IS, or both with COVID-19 severity outcomes stratified by race and gender

We stratified our cohort patients by race and gender to evaluate whether relationships between AID, IS, or AID-IS with COVID-19 severity outcome were similar in the different race and gender groups. Multivariate models, adjusted for demographics and comorbidities, demonstrated that associations of AID, IS, and AID-IS with life-threatening disease were similar in Whites (AID OR 1.12, 95% CI 1.08 -1.16, *P* = 2.34E-9; IS OR 1.27, 95% CI 1.23-1.31, *P* = 5.50E-54 and AID+IS OR 1.33, 95% CI 1.27-1.39, *P* = 1.04E-31) and Black or African Americans (AID OR 1.16, 95% CI 1.06-1.26, *P* = 6.92E-04; IS OR 1.25, 95% CI 1.18-1.33, *P* = 3.24E-14 and AID+IS OR 1.39, 95% CI 1.27-1.52, *P* = 3.60E-13). Similar results were obtained when evaluating hospitalization as an outcome for Whites (AID OR 1.19, 95% CI 1.17-1.22, *P* = 7.10E-61; IS OR 1.20, 95% CI 1.18-1.22, *P* = 1.53E-87 and AID+OR 1.29, 95% CI 1.25-1.33, *P* = 6.19E-65) and Black African Americans (AID OR 1.26, 95% CI 1.21-1.32, *P* = 6.37E-26; IS OR 1.13, 95% CI 1.10-1.17, p = 7.53E-15 and AID+IS OR 1.31, 95% CI 1.24-1.38, *P* = 1.27E-23) (see eTable 4 in the **Supplement** for details).

Stratified analyses by gender showed analogous results, where individuals with a prior diagnosis of AID, exposure to IS or both were more likely to have life-threatening disease and hospitalization in both males and females. Specifically, males and females with prior AID (male OR 1.10, 95% CI 1.04-1.15, *P* = 2.63E-04; female OR 1.16, 95% CI 1.11-1.21, *P* = 5.66E-11, respectively), IS (male OR 1.26, 95% CI 1.22-1.31, *P* = 4.65E-40; female OR 1.27, 95% CI 1.22-1.32, *P* = 2.82E-36, respectively) and AID + IS (male OR 1.33, 95% CI 1.26-1.42, *P* = 6.04E-21; female OR 1.34, 95% CI 1.27-1.41, *P* = 1.59E-27, respectively) are more likely to have life-threatening disease. Similar results were observed when hospitalization was assessed in males (AID OR 1.21, 95% CI 1.17-1.24, *P* = 2.56E-38; IS OR 1.23, 95% CI 1.20-1.25, *P* = 2.35E-82 and AID-IS OR 1.34, 95% CI 1.29-1.39, *P* = 3.24E-49) and females (AID OR 1.21, 95% CI 1.19-1.24, *P* = 3.91E-61; IS OR 1.15, 95% CI 1.13-1.18, *P* = 1.44E-44 and AID-IS OR 1.29, 95% CI 1.25-1.33, *P* = 2.37E-60) respectively (eTable 5 in the **Supplement**).

### Association between AID, IS, or both with COVID-19 severity outcomes in a cohort subset adjusting for COVID-19 vaccination and antiviral exposure

The FDA has previously authorized certain antiviral medications and monoclonal antibodies directed at SARS-CoV-2 to treat mild to moderate COVID-19 in outpatients diagnosed with COVID-19 who were prone to have severe disease manifestations. We specifically considered two small molecule antivirals (Paxlovid(Nirmatrelvir/ritonavir), Lagevrio(molnupiravir), and one monoclonal antibody Bebtelovimab), granted emergency use authorization by the FDA in or after December 2021 (the beginning of the Omicron epoch). Although bebtelovimab has since lost activity against the most recent dominant Omicron variants in the United States (BQ.1,BQ.1.1, and XBB) it was active against prior common US Omicron variants during the period of this study. We evaluated 248,743 patients diagnosed with COVID-19 between December 23rd, 2021 to June 30th 2022, of which 134,812 (54.2%) are vaccinated and 3,974 (1.6%) are exposed to antivirals (see **Methods**, eFigure1c in the **Supplement**). As expected, when adjusting for demographics and comorbidities, we found that usage of antivirals was protective (life-threatening disease: OR 0.31, 95% CI 0.21-0.45; *P* = 4.26E-10 and hospitalization: OR 0.30, 95% CI 0.25-0.36;*P* = 1.39E-40) (eTable 6 in the **Supplement**). Most importantly, independent of exposure to antivirals and vaccination status, hospitalization with COVID-19 was more likely among patients with pre-existing AID (OR= 1.34, 95% CI 1.25 - 1.43, *P* = 8.60E-18), prior IS exposure (OR= 1.61, 95% CI 1.51 - 1.72, *P* = 2.08E-50), or both AID+IS (OR= 1.90, 95% CI 1.73 - 2.10, *P* = 2.39E-39). Similarly, patients with pre-existing AID (OR= 1.18, 95% CI 1.02 - 1.36, *P* = 2.49E-02), prior IS exposure (OR= 1.60, 95% CI 1.42 - 1.81, *P* = 4.71E-14), or both AID + IS (OR= 1.94, 95% CI 1.63 - 2.30, *P* = 1.35E-13) are more likely to have life-threatening disease, independent of exposure to antivirals and vaccination status.

### Association between TNF inhibitors and other IS with COVID-19 severity outcomes in AID patients

In view of prior published data suggesting that TNF inhibitors may be protective against COVID-19^14^, we investigated the association of exposure to TNF inhibitors prior to COVID-19 diagnosis with COVID-19 severity outcomes in patients with a prior AID diagnosis. Of the 191,520 patients with a pre-existing AID diagnosis, 4,789 were exposed to TNF inhibitors at least 14 days prior to COVID-19 diagnosis. When adjusting for demographics and comorbidities, we found that exposure to TNF inhibitors protected against severe COVID-19 outcomes (OR of life-threatening disease 0.80, 95% CI 0.66-0.96, *P* = 1.66E-02), and hospitalization (OR 0.80, 95% CI 0.73-0.89, *P* = 1.06E-05) (**Table3**). While other IS were individually evaluated, only TNF inhibitors showed this protective effect.

**Table 3:** Logistic Regression of severity outcomes in AID patients only with each IS exposure evaluated individually.

|  | Life-threatening condition<br>(Yes: 8,597, No: 182,923)<br>OR (95% CI) | P value | Hospitalized condition<br>(Yes: 30,834, No: 160,686)<br>OR (95% CI) | P value |
| --- | --- | --- | --- | --- |
| **Demographics** |  |  |  |  |
| **Age** | 2.09 (2.02- 2.16) | <2.2E-308 | 1.55 (1.53 - 1.58) | <2.2E-308 |
| **BMI** | 1.04 (1.03 - 1.06) | 3.53E-06 | 1.06 (1.05 - 1.07) | 5.08E-25 |
| **Gender** |  |  |  |  |
| Female | 1 [Reference] |  | 1 [Reference] |  |
| Male | 1.36 (1.30- 1.42) | 1.14E-36 | 1.29 (1.26 - 1.33) | 1.59E-72 |
| **Race** |  |  |  |  |
| White | 1 [Reference] |  | 1 [Reference] |  |
| Others/unknown | 1.13 (1.02- 1.25) | 1.62E-02 | 1.15 (1.09 - 1.22) | 1.04E-06 |
| Black or African American | 1.25 (1.17- 1.34) | 8.05E-12 | 1.63 (1.57 - 1.68) | 1.21E-152 |
| Race asian | 1.26 (1.06- 1.50) | 8.72E-03 | 1.13 (1.03 - 1.25) | 1.39E-02 |
| **Ethnicity** |  |  |  |  |
| Not Hispanic or Latino | 1 [Reference] |  | 1 [Reference] |  |
| Hispanic or Latino | 1.13 (1.03- 1.25) | 1.39E-02 | 1.28 (1.22 - 1.35) | 5.24E-20 |
| **Smoking status** |  |  |  |  |
| Nonsmoker | 1 [Reference] |  | 1 [Reference] |  |
| Current or Former | 1.20 (1.13- 1.27) | 1.89E-10 | 1.39 (1.35 - 1.44) | 1.32E-88 |
| **Comorbidities** |  |  |  |  |
| Cardiovascular disease | 1.88 (1.78- 1.98) | 4.42E-118 | 1.67 (1.62 - 1.72) | 3.13E-243 |
| Dementia | 2.20 (2.04- 2.38) | 7.67E-88 | 2.09 (1.97 - 2.23) | 1.79E-119 |
| Chronic pulmonary disease | 1.24 (1.18- 1.31) | 3.01E-18 | 1.23 (1.20 - 1.27) | 5.36E-46 |
| Liver mild | 1.19 (1.11- 1.27) | 2.02E-07 | 1.13 (1.09 - 1.18) | 4.99E-10 |
| Liver severe | 2.39(2.15 - 2.66) | 2.93E-57 | 1.93 (1.78 - 2.09) | 5.40E-60 |
| Kidney disease | 1.94 (1.85- 2.05) | 3.49E-139 | 1.89 (1.83 - 1.95) | <2.2E-308 |
| Cancer | 1.32 (1.25- 1.40) | 6.41E-20 | 1.16 (1.12 - 1.21) | 1.09E-13 |
| Metastatic cancer | 2.36 (2.12- 2.62) | 1.94E-56 | 1.67 (1.54 - 1.81) | 1.66E-33 |
| Type 2 Diabetes Mellitus | 1.35 (1.29- 1.42) | 1.58E-32 | 1.50 (1.46 - 1.55) | 6.22E-167 |
| HIV | 1.00 (0.77 - 1.30) | 9.94E-01 | 1.05 (0.90 - 1.22) | 5.57E-01 |
| **Immunosuppressants** |  |  |  |  |
| TNF inhibitor | 0.80 (0.66 - 0.96) | 1.66E-02 | 0.80 (0.73 - 0.89) | 1.06E-05 |
| Calcineurin inhibitor | 0.95(0.84 - 1.08) | 4.61E-01 | 0.99 (0.91 - 1.07) | 6.99E-01 |
| IL inhibitor | 1.03 (0.84 - 1.27) | 7.59E-01 | 1.08 (0.96 - 1.21) | 2.21E-01 |
| Other selective IS | 1.04 (0.91 - 1.20) | 5.48E-01 | 1.15 (1.06 - 1.25) | 5.95E-04 |
| Cyclophosphamide | 1.05 (0.77 - 1.44) | 7.39E-01 | 0.90 (0.72 - 1.12) | 3.47E-01 |
| Glucocorticoid | 1.12 (1.06- 1.18) | 5.51E-05 | 1.04 (1.01 - 1.08) | 1.30E-02 |
| Azathioprine | 1.13 (0.93- 1.38) | 2.22E-01 | 1.17 (1.04 - 1.31) | 8.02E-03 |
| Other antineoplastic agents | 1.24 (1.12- 1.37) | 4.33E-05 | 1.04 (0.98 - 1.11) | 2.22E-01 |
| JAK inhibitor | 1.28 (0.91- 1.79) | 1.59E-01 | 1.37 (1.14 - 1.66) | 1.11E-03 |
| Monoclonal other | 1.33 (1.07- 1.65) | 1.07E-02 | 1.09 (0.92 - 1.29) | 3.03E-01 |
| Mycophenol | 1.44 (1.25- 1.65) | 1.91E-07 | 1.60 (1.47 - 1.74) | 6.64E-28 |
| Anthracyclines | 1.54 (1.10- 2.14) | 1.12E-02 | 1.53 (1.19 - 1.98) | 1.16E-03 |
| Rituximab | 1.62 (1.35- 1.95) | 3.16E-07 | 1.73 (1.53 - 1.96) | 1.65E-18 |
| PK inhibitor | 1.63 (1.32- 2.01) | 5.91E-06 | 1.76 (1.50 - 2.08) | 9.48E-12 |
| Checkpoint inhibitor | 1.70 (1.28- 2.26) | 2.72E-04 | 1.29 (0.99 - 1.67) | 5.81E-02 |

Demographic factors include age, BMI, gender, race, ethnicity, smoking status.
Comorbidities include cardiovascular disease (MI + CHF + PVD + stroke), dementia, pulmonary disease, liver disease (mild and severe), Type 2 diabetes, kidney disease, cancer (metastatic and non-metastatic), and HIV infection.
Immunosuppressants: Anthracyclines, Checkpoint inhibitor, Cyclophosphamide, PK inhibitor, Monoclonal other, Rituximab, Other antineoplastic agents, Azathioprine, Calcineurin inhibitor, IL inhibitor, JAK inhibitor, Mycophenol, TNF\_inhibitor, Other selective immunosuppressants, Glucocorticoid.

## Discussion

Using N3C, the largest publicly available clinical data set of US-based patients diagnosed with COVID-19, we identified 2,453,799 patients diagnosed with PCR or antigen testing-confirmed COVID-19, of whom 191,520 had a prior diagnosis of AID, 278,095 had a prior exposure to long-term immunosuppressants (IS), and 56,813 had both a prior AID diagnosis and exposure to IS. Rheumatoid arthritis was the most common AID. Our cohort is based on data from the beginning of the pandemic to June 30^th^ 2022, which includes epochs spanning the ancestral strain as well as five major variants (Alpha, Beta, Gamma, Delta, and Omicron). Overall, this cohort thus appropriately represent a broad population of patients diagnosed with COVID-19 and AID in the US over time.

We found that COVID-19 patients that have had a prior diagnosis of AID, exposure to IS, or both a prior AID diagnosis and IS exposure are more likely to have life-threatening disease or to be hospitalized. These results were robust as they were adjusted for basic demographics and comorbidities, and confirm that each and both prior AID diagnosis and exposure to IS are risk factors for worse COVID-19 disease outcomes. Further, we performed a sensitivity analysis in a subset of our cohort with available vaccination and antiviral exposure data (December 23rd 2021 to June 30th 2022) and confirmed that AID and/or IS are risk factors of worse COVID-19 outcomes, independent of whether patients are exposed to antiviral treatments (nirmatrelvir/ritonavir, LAGEVRIO (molnupiravir), and bebtelovimab) and/or had at least one COVID-19 vaccination dose. Our results thus help clarify the ambiguity in previous studies in answering the difficult question of whether prior AID diagnosis or exposure to IS are risk factors for worse COVID-19 disease outcomes.

Race and sex are known to be associated with COVID-19 severity. A recent study of race and ethnicity-based COVID-19 outcome disparity in the United States population reported that Asian American individuals had a higher risk of COVID-19 positivity and ICU admission than White individuals^24,25.^ Further, socioeconomic disparity and clinical care quality are associated with COVID-19 mortality and incidence in racial and ethnic minority groups^25^. A recent study has indicated that the severity and mortality of COVID-19 is higher in males than in females^26,27^. To ensure that our results were robust to differences in race and sex, we stratified our evaluations by race and sex to evaluate effects of AID, IS, or AID and IS exposure on COVID-19 severity outcomes in these various groups. Our results indicate that our findings broadly apply to race and both genders.

Finally, our study design enabled us to further clarify whether some specific IS showed contrary effects. Indeed, a recent study suggested TNF inhibitor monotherapy was associated with a lower risk of adverse COVID-19 outcomes compared to other commonly prescribed immunotherapy among patients with AID^29^. In this study, we confirm that patients with a prior AID diagnosis and exposed to TNF inhibitors prior to infection are less likely to be hospitalized or have life-threatening COVID-19. We also confirmed that this protective effect is unique to TNF inhibitors, and that other IS are associated with either higher or no risk of worse COVID-19 severity outcomes.

There are limitations to this study worth noting. First, the medical history of COVID-19 patients is limited to January 1, 2018 or later, with some patients having limited interaction with participating health care systems prior to their index diagnosis, making it difficult to fully assess pre-existing conditions and comorbidities. The diagnosis date of AID is thus challenging to determine precisely. To mitigate these risks, patients with at least one encounter visit before diagnosis were considered to increase the robustness of past medical history documentation. Further, we note that N3C data are aggregated from many health care systems, covering four common data models that vary in granularity. Harmonization of these disparate data thus requires assumptions and inferences to be made that could incur systematic biases. Similarly, the ability to accurately determine race within N3C is diminished by variations in how race is reported in different healthcare systems^30^ Nonetheless, we highlight the meticulous efforts from the N3C collaborative in evaluating and improving the quality of phenotypes generated within^31^ N3C. Further, missingness is a known issue with N3C’s vaccination data, as patients may receive vaccine doses at pop-up clinics, drugstores, or at their place of employment, which may not end up recorded in the patient’s records. To counteract this missingness in our vaccination subanalysis, we only included sites whose rate of vaccination in N3C data was within range of the CDC’s vaccination rate for that site’s geographic region^32^. Finally, we also recognize limitations related to the retrospective design of this study, inability to handle all possible confounders, and the possibility that follow-up data among patients could be incomplete (e.g. patients could have sought care in institutions that are not affiliated with N3C). Despite these limitations, this study is an important step toward increasing our understanding, at a population level, of whether prior exposure to AID, IS, or both pose an additional risk to patients in developing worse COVID-19 disease outcomes.

## Conclusions

To the best of our knowledge, this study represents the largest, most comprehensive systematic analysis of the effects of AID, IS, or both on COVID-19 severity outcomes. Our study suggests that patients with a prior AID diagnosis, prior exposure to IS or both AID and IS have a higher risk of life-threatening COVID-19 disease or hospitalization. These associations were consistent in different race subsets (African Americans, Whites) and gender subsets. Importantly, these results provide a more definitive answer to previous discrepant findings on whether patients with AID are at higher risk for worse COVID-19 related outcomes, providing clinicians with helpful data that may help guide their treatment and monitoring plans.

## Data Availability

The analyses described in this publication were conducted with data or tools accessed through the
NCATS N3C Data Enclave covid.cd2h.org/enclave and supported by CD2H - The National COVID Cohort
Collaborative (N3C) IDeA CTR Collaboration 3U24TR002306-04S2 NCATS U24 TR002306. This research
was possible because of the patients whose information is included within the data from participating
organizations (covid.cd2h.org/dtas) and the organizations and scientists (covid.cd2h.org/duas) who have
contributed to the on-going development of this community resource. Enclave data is protected, and
can be accessed for COVID-related research with an approved (1) IRB protocol and (2) Data Use Request
(DUR). Enclave and data access instructions can be found at https://covid.cd2h.org/for-researchers; all
code used to produce the analyses in this manuscript is available within the N3C Enclave to users with
valid login credentials to support reproducibility

## Acknowledgements

This work was supported in part by the Intramural and Extramural Research Program of the National Center for Advancing Translational Sciences, National Institutes of Health (ZICTR000410-03), and in part by the Intramural Research Programs of the National Institute of Diabetes and Digestive and Kidney Diseases (project number ZIA/DK075149 to BA). We would like to thank Sam Michael and Kenneth Gersing for their help and support in getting access to the N3C data enclave seamlessly.

## N3C Acknowledgements

The code for this study (DUR: RP-42D046) is located within the N3C Data Enclave and can be accessed upon request, provided the user already has access to the N3C. Authorship was determined using ICMJE recommendations.

### N3C Attribution

The analyses described in this publication were conducted with data or tools accessed through the NCATS N3C Data Enclave covid.cd2h.org/enclave and supported by CD2H - The National COVID Cohort Collaborative (N3C) IDeA CTR Collaboration 3U24TR002306-04S2 NCATS U24 TR002306. This research was possible because of the patients whose information is included within the data from participating organizations (covid.cd2h.org/dtas) and the organizations and scientists (covid.cd2h.org/duas) who have contributed to the on-going development of this community resource (cite this https://doi.org/10.1093/jamia/ocaa196).

### Disclaimer

The content is solely the responsibility of the authors and does not necessarily represent the official views of the National Institutes of Health or the N3C program.

### IRB

The N3C data transfer to NCATS is performed under a Johns Hopkins University Reliance Protocol # IRB00249128 or individual site agreements with NIH. The N3C Data Enclave is managed under the authority of the NIH; information can be found at https://ncats.nih.gov/n3c/resources.

### Individual Acknowledgements for Core Contributors

We gratefully acknowledge the following core contributors to N3C:

Adam B. Wilcox, Adam M. Lee, Alexis Graves, Alfred (Jerrod) Anzalone, Amin Manna, Amit Saha, Amy Olex, Andrea Zhou, Andrew E. Williams, Andrew Southerland, Andrew T. Girvin, Anita Walden, Anjali A. Sharathkumar, Benjamin Amor, Benjamin Bates, Brian Hendricks, Brijesh Patel, Caleb Alexander, Carolyn Bramante, Cavin Ward-Caviness, Charisse Madlock-Brown, Christine Suver, Christopher Chute, Christopher Dillon, Chunlei Wu, Clare Schmitt, Cliff Takemoto, Dan Housman, Davera Gabriel, David A. Eichmann, Diego Mazzotti, Don Brown, Eilis Boudreau, Elaine Hill, Elizabeth Zampino, Emily Carlson Marti, Emily R. Pfaff, Evan French, Farrukh M Koraishy, Federico Mariona, Fred Prior, George Sokos, Greg Martin, Harold Lehmann, Heidi Spratt, Hemalkumar Mehta, Hongfang Liu, Hythem Sidky, J.W. Awori Hayanga, Jami Pincavitch, Jaylyn Clark, Jeremy Richard Harper, Jessica Islam, Jin Ge, Joel Gagnier, Joel H. Saltz, Joel Saltz, Johanna Loomba, John Buse, Jomol Mathew, Joni L. Rutter, Julie A. McMurry, Justin Guinney, Justin Starren, Karen Crowley, Katie Rebecca Bradwell, Kellie M. Walters, Ken Wilkins, Kenneth R. Gersing, Kenrick Dwain Cato, Kimberly Murray, Kristin Kostka, Lavance Northington, Lee Allan Pyles, Leonie Misquitta, Lesley Cottrell, Lili Portilla, Mariam Deacy, Mark M. Bissell, Marshall Clark, Mary Emmett, Mary Morrison Saltz, Matvey B. Palchuk, Melissa A. Haendel, Meredith Adams, Meredith Temple-O’Connor, Michael G. Kurilla, Michele Morris, Nabeel Qureshi, Nasia Safdar, Nicole Garbarini, Noha Sharafeldin, Ofer Sadan, Patricia A. Francis, Penny Wung Burgoon, Peter Robinson, Philip R.O. Payne, Rafael Fuentes, Randeep Jawa, Rebecca Erwin-Cohen, Rena Patel, Richard A. Moffitt, Richard L. Zhu, Rishi Kamaleswaran, Robert Hurley, Robert T. Miller, Saiju Pyarajan, Sam G. Michael, Samuel Bozzette, Sandeep Mallipattu, Satyanarayana Vedula, Scott Chapman, Shawn T. O’Neil, Soko Setoguchi, Stephanie S. Hong, Steve Johnson, Tellen D. Bennett, Tiffany Callahan, Umit Topaloglu, Usman Sheikh, Valery Gordon, Vignesh Subbian, Warren A. Kibbe, Wenndy Hernandez, Will Beasley, Will Cooper, William Hillegass, Xiaohan Tanner Zhang. Details of contributions available at covid.cd2h.org/core-contributors

### Data Partners with Released Data

The following institutions whose data is released or pending:

#### Available

Advocate Health Care Network — UL1TR002389: The Institute for Translational Medicine (ITM) • Boston University Medical Campus — UL1TR001430: Boston University Clinical and Translational Science Institute • Brown University — U54GM115677: Advance Clinical Translational Research (Advance-CTR) • Carilion Clinic — UL1TR003015: iTHRIV Integrated Translational health Research Institute of Virginia • Charleston Area Medical Center — U54GM104942: West Virginia Clinical and Translational Science Institute (WVCTSI) • Children’s Hospital Colorado — UL1TR002535: Colorado Clinical and Translational Sciences Institute • Columbia University Irving Medical Center — UL1TR001873: Irving Institute for Clinical and Translational Research • Duke University — UL1TR002553: Duke Clinical and Translational Science Institute • George Washington Children’s Research Institute — UL1TR001876: Clinical and Translational Science Institute at Children’s National (CTSA-CN) • George Washington University — UL1TR001876: Clinical and Translational Science Institute at Children’s National (CTSA-CN) • Indiana University School of Medicine — UL1TR002529: Indiana Clinical and Translational Science Institute • Johns Hopkins University — UL1TR003098: Johns Hopkins Institute for Clinical and Translational Research • Loyola Medicine — Loyola University Medical Center • Loyola University Medical Center — UL1TR002389: The Institute for Translational Medicine (ITM) • Maine Medical Center — U54GM115516: Northern New England Clinical & Translational Research (NNE-CTR) Network • Massachusetts General Brigham — UL1TR002541: Harvard Catalyst • Mayo Clinic Rochester — UL1TR002377: Mayo Clinic Center for Clinical and Translational Science (CCaTS) • Medical University of South Carolina — UL1TR001450: South Carolina Clinical & Translational Research Institute (SCTR) • Montefiore Medical Center — UL1TR002556: Institute for Clinical and Translational Research at Einstein and Montefiore • Nemours — U54GM104941: Delaware CTR ACCEL Program • NorthShore University HealthSystem — UL1TR002389: The Institute for Translational Medicine (ITM) • Northwestern University at Chicago — UL1TR001422: Northwestern University Clinical and Translational Science Institute (NUCATS) • OCHIN — INV-018455: Bill and Melinda Gates Foundation grant to Sage Bionetworks • Oregon Health & Science University — UL1TR002369: Oregon Clinical and Translational Research Institute • Penn State Health Milton S. Hershey Medical Center — UL1TR002014: Penn State Clinical and Translational Science Institute • Rush University Medical Center — UL1TR002389: The Institute for Translational Medicine (ITM) • Rutgers, The State University of New Jersey — UL1TR003017: New Jersey Alliance for Clinical and Translational Science • Stony Brook University — U24TR002306 • The Ohio State University — UL1TR002733: Center for Clinical and Translational Science • The State University of New York at Buffalo — UL1TR001412: Clinical and Translational Science Institute • The University of Chicago — UL1TR002389: The Institute for Translational Medicine (ITM) • The University of Iowa — UL1TR002537: Institute for Clinical and Translational Science • The University of Miami Leonard M. Miller School of Medicine — UL1TR002736: University of Miami Clinical and Translational Science Institute • The University of Michigan at Ann Arbor — UL1TR002240: Michigan Institute for Clinical and Health Research • The University of Texas Health Science Center at Houston — UL1TR003167: Center for Clinical and Translational Sciences (CCTS) • The University of Texas Medical Branch at Galveston — UL1TR001439: The Institute for Translational Sciences • The University of Utah — UL1TR002538: Uhealth Center for Clinical and Translational Science • Tufts Medical Center — UL1TR002544: Tufts Clinical and Translational Science Institute • Tulane University — UL1TR003096: Center for Clinical and Translational Science • University Medical Center New Orleans — U54GM104940: Louisiana Clinical and Translational Science (LA CaTS) Center • University of Alabama at Birmingham — UL1TR003096: Center for Clinical and Translational Science • University of Arkansas for Medical Sciences — UL1TR003107: UAMS Translational Research Institute • University of Cincinnati — UL1TR001425: Center for Clinical and Translational Science and Training • University of Colorado Denver, Anschutz Medical Campus — UL1TR002535: Colorado Clinical and Translational Sciences Institute • University of Illinois at Chicago — UL1TR002003: UIC Center for Clinical and Translational Science • University of Kansas Medical Center — UL1TR002366: Frontiers: University of Kansas Clinical and Translational Science Institute • University of Kentucky — UL1TR001998: UK Center for Clinical and Translational Science • University of Massachusetts Medical School Worcester — UL1TR001453: The UMass Center for Clinical and Translational Science (UMCCTS) • University of Minnesota — UL1TR002494: Clinical and Translational Science Institute • University of Mississippi Medical Center — U54GM115428: Mississippi Center for Clinical and Translational Research (CCTR) • University of Nebraska Medical Center — U54GM115458: Great Plains IDeA-Clinical & Translational Research • University of North Carolina at Chapel Hill — UL1TR002489: North Carolina Translational and Clinical Science Institute • University of Oklahoma Health Sciences Center — U54GM104938: Oklahoma Clinical and Translational Science Institute (OCTSI) • University of Rochester — UL1TR002001: UR Clinical & Translational Science Institute • University of Southern California — UL1TR001855: The Southern California Clinical and Translational Science Institute (SC CTSI) • University of Vermont — U54GM115516: Northern New England Clinical & Translational Research (NNE-CTR) Network • University of Virginia — UL1TR003015: iTHRIV Integrated Translational health Research Institute of Virginia • University of Washington — UL1TR002319: Institute of Translational Health Sciences • University of Wisconsin-Madison — UL1TR002373: UW Institute for Clinical and Translational Research • Vanderbilt University Medical Center — UL1TR002243: Vanderbilt Institute for Clinical and Translational Research • Virginia Commonwealth University — UL1TR002649: C. Kenneth and Dianne Wright Center for Clinical and Translational Research • Wake Forest University Health Sciences — UL1TR001420: Wake Forest Clinical and Translational Science Institute • Washington University in St. Louis — UL1TR002345: Institute of Clinical and Translational Sciences • Weill Medical College of Cornell University — UL1TR002384: Weill Cornell Medicine Clinical and Translational Science Center • West Virginia University — U54GM104942: West Virginia Clinical and Translational Science Institute (WVCTSI)

#### Submitted

Icahn School of Medicine at Mount Sinai — UL1TR001433: ConduITS Institute for Translational Sciences • The University of Texas Health Science Center at Tyler — UL1TR003167: Center for Clinical and Translational Sciences (CCTS) • University of California, Davis — UL1TR001860: UCDavis Health Clinical and Translational Science Center • University of California, Irvine — UL1TR001414: The UC Irvine Institute for Clinical and Translational Science (ICTS) • University of California, Los Angeles — UL1TR001881: UCLA Clinical Translational Science Institute • University of California, San Diego — UL1TR001442: Altman Clinical and Translational Research Institute • University of California, San Francisco — UL1TR001872: UCSF Clinical and Translational Science Institute

#### Pending

Arkansas Children’s Hospital — UL1TR003107: UAMS Translational Research Institute • Baylor College of Medicine — None (Voluntary) • Children’s Hospital of Philadelphia — UL1TR001878: Institute for Translational Medicine and Therapeutics • Cincinnati Children’s Hospital Medical Center — UL1TR001425: Center for Clinical and Translational Science and Training • Emory University — UL1TR002378: Georgia Clinical and Translational Science Alliance • HonorHealth — None (Voluntary) • Loyola University Chicago — UL1TR002389: The Institute for Translational Medicine (ITM) • Medical College of Wisconsin — UL1TR001436: Clinical and Translational Science Institute of Southeast Wisconsin • MedStar Health Research Institute — UL1TR001409: The Georgetown-Howard Universities Center for Clinical and Translational Science (GHUCCTS) • MetroHealth — None (Voluntary) • Montana State University — U54GM115371: American Indian/Alaska Native CTR • NYU Langone Medical Center — UL1TR001445: Langone Health’s Clinical and Translational Science Institute • Ochsner Medical Center — U54GM104940: Louisiana Clinical and Translational Science (LA CaTS) Center • Regenstrief Institute — UL1TR002529: Indiana Clinical and Translational Science Institute • Sanford Research — None (Voluntary) • Stanford University — UL1TR003142: Spectrum: The Stanford Center for Clinical and Translational Research and Education • The Rockefeller University — UL1TR001866: Center for Clinical and Translational Science • The Scripps Research Institute — UL1TR002550: Scripps Research Translational Institute • University of Florida — UL1TR001427: UF Clinical and Translational Science Institute • University of New Mexico Health Sciences Center — UL1TR001449: University of New Mexico Clinical and Translational Science Center • University of Texas Health Science Center at San Antonio — UL1TR002645: Institute for Integration of Medicine and Science • Yale New Haven Hospital — UL1TR001863: Yale Center for Clinical Investigation

**For the source code** see:

“https://github.com/arjunyadaw/N3C-Autoimmune-Disease-model.git“

## Author Contributions

*Concept and design: ASY, BA, DS, EAM*

*Acquisition, analysis, or interpretation of data: All authors*

*Drafting of the manuscript: ASY and EAM*

*Critical revision of the manuscript for important intellectual content: All authors*

*Statistical analysis: ASY, EAM, and HS*

*Obtained funding: EAM*

*Administrative, technical, or material support: NH and HS*

*Supervision: EAM*

## Bibliography

1. Johns Hopkins University & Medicine. Coronavirus Research Center. COVID-19 dashboard. Johns Hopkins University. 2021. Accessed January 3, 2022. https://coronavirus.jhu.edu/map.html

2. Coronavirus disease (COVID-19): Post COVID-19 condition. https://www.who.int/news-room/questions-and-answers/item/coronavirus-disease-(covid-19)-post-covid-19-condition

3. CDC. People with Certain Medical Conditions. Centers for Disease Control and Prevention. https://www.cdc.gov/coronavirus/2019-ncov/need-extra-precautions/people-with-medical-conditions.html

4. NIH Autoimmune Diseases Coordinating Committee: Autoimmune Diseases Research Plan, March 2005. https://www.niaid.nih.gov/sites/default/files/adccfinal.pdf

5. Serling-Boyd N, D’Silva KM, Hsu TY, et al. Coronavirus disease 2019 outcomes among patients with rheumatic diseases 6 months into the pandemic. Ann Rheum Dis. 2021;80(5):660–666. doi:10.1136/annrheumdis-2020-219279

6. D’Silva KM, Serling-Boyd N, Wallwork R, et al. Clinical characteristics and outcomes of patients with coronavirus disease 2019 (COVID-19) and rheumatic disease: a comparative cohort study from a US “hot spot.” Ann Rheum Dis. 2020;79(9):1156–1162. doi:10.1136/annrheumdis-2020-217888

7. Ye C, Cai S, Shen G, et al. Clinical features of rheumatic patients infected with COVID-19 in Wuhan, China. Ann Rheum Dis. 2020;79(8):1007–1013. doi:10.1136/annrheumdis-2020-217627

8. Rodríguez-Lago I, Ramírez de la Piscina P, Elorza A, Merino O, Ortiz de Zárate J, Cabriada JL. Characteristics and Prognosis of Patients With Inflammatory Bowel Disease During the SARS-CoV-2 Pandemic in the Basque Country (Spain). Gastroenterology. 2020;159(2):781–783. doi:10.1053/j.gastro.2020.04.043

9. Faye AS, Lee KE, Laszkowska M, et al. Risk of Adverse Outcomes in Hospitalized Patients with Autoimmune Disease and COVID-19: A Matched Cohort Study from New York City. J Rheumatol. 2021;48(3):454–462. doi:10.3899/jrheum.200989

10. Akiyama S, Hamdeh S, Micic D, Sakuraba A. Prevalence and clinical outcomes of COVID-19 in patients with autoimmune diseases: a systematic review and meta-analysis. Ann Rheum Dis. 2021;80(3):384–391. doi:10.1136/annrheumdis-2020-218946

11. Pereira MR, Mohan S, Cohen DJ, et al. COVID-19 in solid organ transplant recipients: Initial report from the US epicenter. Am J Transplant. 2020;20(7):1800–1808. doi:10.1111/ajt.15941

12. Fung M, Babik JM. COVID-19 in Immunocompromised Hosts: What We Know So Far. Clin Infect Dis Off Publ Infect Dis Soc Am. 2021;72(2):340–350. doi:10.1093/cid/ciaa863

13. Andersen KM, Bates BA, Rashidi ES, et al. Long-term use of immunosuppressive medicines and in-hospital COVID-19 outcomes: a retrospective cohort study using data from the National COVID Cohort Collaborative. Lancet Rheumatol. 2022;4(1):e33–e41. doi:10.1016/S2665-9913(21)00325-8

14. Kridin K, Schonmann Y, Damiani G, et al. Tumor necrosis factor inhibitors are associated with a decreased risk of COVID -19-associated hospitalization in patients with psoriasis—A population-based cohort study. Dermatol Ther. 2021;34(4). doi:10.1111/dth.15003

15. Immune modulator drugs improved survival for people hospitalized with COVID-19. https://www.nih.gov/news-events/news-releases/immune-modulator-drugs-improved-survival-people-hospitalized-covid-19.

16. Bennett TD, Moffitt RA, Hajagos JG, et al. Clinical Characterization and Prediction of Clinical Severity of SARS-CoV-2 Infection Among US Adults Using Data From the US National COVID Cohort Collaborative. JAMA Netw Open. 2021;4(7):e2116901. doi:10.1001/jamanetworkopen.2021.16901

17. Haendel MA, Chute CG, Bennett TD, et al. The National COVID Cohort Collaborative (N3C): Rationale, design, infrastructure, and deployment. J Am Med Inform Assoc JAMIA. 2021;28(3):427–443. doi:10.1093/jamia/ocaa196

18. GitHub. National COVID Cohort Collaborative, COVID-19 Phenotype Documentation, version 4.0 (last updated 3/11/2022). Accessed September 30, 2021. https://github.com/National-COVID-Cohort-Collaborative/Phenotype_Data_Acquisition/wiki/Latest-Phenotype

19. Sun J, Zheng Q, Madhira V, et al. Association Between Immune Dysfunction and COVID-19 Breakthrough Infection After SARS-CoV-2 Vaccination in the US. JAMA Intern Med. 2022;182(2):153. doi:10.1001/jamainternmed.2021.7024

20. Marshall JC, Murthy S, Diaz J, et al. A minimal common outcome measure set for COVID-19 clinical research. Lancet Infect Dis. 2020;20(8):e192–e197. doi:10.1016/S1473-3099(20)30483-7

21. GAI. Autoimmune Disease List. Global Autoimmune Institute. Accessed August 12, 2022. https://www.autoimmuneinstitute.org/resources/autoimmune-disease-list/

22. Autoimmune disease. Accessed August 12, 2022. https://www.ebi.ac.uk/ols/ontologies/EFO/terms?iri=http://www.ebi.ac.uk/efo/EFO_0005140

23. Pollard TJ, Johnson AEW, Raffa JD, Mark RG. tableone: An open source Python package for producing summary statistics for research papers. JAMIA Open. 2018;1(1):26–31. doi:10.1093/jamiaopen/ooy012

24. Fu J, Reid SA, French B, et al. Racial Disparities in COVID-19 Outcomes Among Black and White Patients With Cancer. JAMA Netw Open. 2022;5(3):e224304. doi:10.1001/jamanetworkopen.2022.4304

25. Magesh S, John D, Li WT, et al. Disparities in COVID-19 Outcomes by Race, Ethnicity, and Socioeconomic Status: A Systematic Review and Meta-analysis. JAMA Netw Open. 2021;4(11):e2134147. doi:10.1001/jamanetworkopen.2021.34147

26. Chaturvedi R, Lui B, Aaronson JA, White RS, Samuels JD. COVID-19 complications in males and females: recent developments. J Comp Eff Res. 2022;11(9):689–698. doi:10.2217/cer-2022-0027

27. Pradhan A, Olsson PE. Sex differences in severity and mortality from COVID-19: are males more vulnerable? Biol Sex Differ. 2020;11(1):53. doi:10.1186/s13293-020-00330-7

28. Conrad N, Verbeke G, Molenberghs G, et al. Autoimmune diseases and cardiovascular risk: a population-based study on 19 autoimmune diseases and 12 cardiovascular diseases in 22 million individuals in the UK. The Lancet. 2022;400(10354):733–743. doi:10.1016/S0140-6736(22)01349-6

29. Izadi Z, Brenner EJ, Mahil SK, et al. Association Between Tumor Necrosis Factor Inhibitors and the Risk of Hospitalization or Death Among Patients With Immune-Mediated Inflammatory Disease and COVID-19. JAMA Netw Open. 2021;4(10):e2129639. doi:10.1001/jamanetworkopen.2021.29639

30. Cook L, Espinoza J, Weiskopf NG, et al. Issues With Variability in Electronic Health Record Data About Race and Ethnicity: Descriptive Analysis of the National COVID Cohort Collaborative Data Enclave. JMIR Med Inform. 2022;10(9):e39235. doi:10.2196/39235

31. Pfaff ER, Girvin AT, Gabriel DL, et al. Synergies between centralized and federated approaches to data quality: a report from the national COVID cohort collaborative. J Am Med Inform Assoc. 2022;29(4):609–618. doi:10.1093/jamia/ocab217

32. Brannock MD, Chew RF, Preiss AJ, et al. Long COVID Risk and Pre-COVID Vaccination: An EHR-Based Cohort Study from the RECOVER Program. Epidemiology; 2022. doi:10.1101/2022.10.06.22280795

