## Supplemental tables and figure for "Pre-existing autoimmunity is associated with increased severity of COVID-19: A retrospective cohort study using data from the National COVID Cohort Collaborative (N3C)"

**eTable 1. Preexisting Comorbidities evaluated with associated concept set names and IDs.**
Preexisting comorbidities were defined as occurring prior to COVID-19 diagnosis.

| Pre-existing Comorbidity | Concept Set Names (latest codeset ID) |
|---|---|
| Myocardial infarction (MI) | Charlson - MI (259495957) |
| Congestive heart failure (CHF) | Charlson - CHF (359043664) |
| Peripheral vascular disease (PVD) | Charlson - PVD (376881697) |
| Stroke | Charlson - Stroke (652711186) |
| Dementia | Charlson - Dementia (78746470) |
| Pulmonary Diseases | Charlson - Pulmonary (514953976) |
| Liver diseases, mild | Charlson - LiverMild (494981955) |
| Liver diseases, severe | Charlson - LiverSevere (248333963) |
| Type 2 diabetes mellitus | Type 2 diabetes mellitus (484742674) |
| Kidney diseases | Charlson - Kidney (220495690) |
| Cancer | Charlson - Cancer (535274723) |
| Metastatic cancer (Mets) | Charlson - Mets (378462283) |
| HIV infection | HIV Infection (382527336) |

**eTable 2. List of autoimmune diseases with its concept Ids**
AID diagnosis cocured prior to the earliest covid diagnosis. AID sorted based on OMOP, Vocabulary Id = SNOMED & Concept Class Id = Clinical Finding from condition_occurence table.

| | Autoimmune Disease Names | Concept ID |
|---|---|---|
| **Type 1 diabetes** | Type 1 diabetes mellitus with unspecified diabetic retinopathy | 1567944 |
| | Type 1 diabetes mellitus with moderate nonproliferative diabetic retinopathy | 1567946 |
| | Type 1 diabetes mellitus with oral complications | 1567954 |
| | Poorly controlled type 1 diabetes | 3194119 |
| | Latent autoimmune diabetes mellitus in adult | 4145827 |

| | |
|---|---|
| Type 1 diabetes mellitus with unspecified complications | 35206878 |
| Mild nonproliferative retinopathy due to type 1 diabetes mellitus | 37016179 |
| Hyperglycemia due to type 1 diabetes mellitus | 37016348 |
| Type 1 diabetes mellitus with mild nonproliferative diabetic retinopathy with macular edema, left eye | 37200142 |
| Type 1 diabetes mellitus with mild nonproliferative diabetic retinopathy with macular edema, unspecified eye | 37200144 |
| Type 1 diabetes mellitus with moderate nonproliferative diabetic retinopathy with macular edema, unspecified eye | 37200152 |
| Type 1 diabetes mellitus with severe nonproliferative diabetic retinopathy with macular edema, right eye | 37200157 |
| Type 1 diabetes mellitus with severe nonproliferative diabetic retinopathy with macular edema, unspecified eye | 37200160 |
| Type 1 diabetes mellitus with severe nonproliferative diabetic retinopathy without macular edema, left eye | 37200162 |
| Type 1 diabetes mellitus with proliferative diabetic retinopathy with macular edema, bilateral | 37200167 |
| Type 1 diabetes mellitus with proliferative diabetic retinopathy with traction retinal detachment involving the macula, bilateral | 37200172 |
| Type 1 diabetes mellitus with proliferative diabetic retinopathy with combined traction retinal detachment and rhegmatogenous retinal detachment, left eye | 37200181 |
| Type 1 diabetes mellitus with stable proliferative diabetic retinopathy, bilateral | 37200187 |
| Disorder of eye due to type 1 diabetes mellitus | 42538169 |
| Pre-existing type 1 diabetes mellitus in pregnancy | 43531008 |
| Type 1 diabetes mellitus with hyperglycemia | 45533018 |
| Type 1 diabetes mellitus with other diabetic arthropathy | 45537960 |
| Type 1 diabetes mellitus with unspecified diabetic retinopathy without macular edema | 45542736 |
| Type 1 diabetes mellitus with diabetic peripheral angiopathy without gangrene | 45547622 |
| Type 1 diabetes mellitus with other oral complications | 45547623 |

| | | |
|---|---|---|
| | Type 1 diabetes mellitus with other diabetic ophthalmic complication | 45576438 |
| | Type 1 diabetes mellitus with mild nonproliferative diabetic retinopathy without macular edema | 45591026 |
| | Type 1 diabetes mellitus with other diabetic kidney complication | 45600637 |
| | Type 1 diabetes mellitus with diabetic dermatitis | 45600639 |
| | Type 1 diabetes mellitus with hypoglycemia without coma | 45600640 |
| | Hypoglycemic unawareness due to type 1 diabetes mellitus | 45757362 |
| | Microalbuminuria due to type 1 diabetes mellitus | 45757535 |
| | Diabetes mellitus type 1 without retinopathy | 45757674 |
| | Blindness due to type 1 diabetes mellitus | 45763585 |
| | Neuropathic arthropathy due to type 1 diabetes mellitus | 45769830 |
| | Severe malnutrition due to type 1 diabetes mellitus | 45769833 |
| | Hypoglycemia due to type 1 diabetes mellitus | 45769876 |
| | Chronic kidney disease stage 1 due to type 1 diabetes mellitus | 45773576 |
| | Diabetes Type 1 | 45883360 |
| | Renal disorder due to type 1 diabetes mellitus | 200687 |
| | Peripheral circulatory disorder due to type 1 diabetes mellitus | 318712 |
| | Ketoacidosis due to type 1 diabetes mellitus | 439770 |
| | Type 1 diabetes mellitus without complication | 443412 |
| | Type 1 diabetes mellitus maturity onset | 4099215 |
| | Lumbosacral radiculoplexus neuropathy due to type 1 diabetes mellitus | 4143857 |
| | Exudative maculopathy due to type 1 diabetes mellitus | 4221344 |
| | Persistent proteinuria due to type 1 diabetes mellitus | 4222553 |
| | Ketoacidotic coma due to type 1 diabetes mellitus | 4224254 |
| | Multiple complications due to type 1 diabetes mellitus | 4224709 |
| | Cataract due to diabetes mellitus type 1 | 4225656 |
| | Type 1 diabetes mellitus with hypoglycemia without coma | 35206877 |
| | Moderate nonproliferative retinopathy due to type 1 diabetes mellitus | 37016180 |

| | |
|---|---|
| Type 1 diabetes mellitus with mild nonproliferative diabetic retinopathy with macular edema, bilateral | 37200143 |
| Type 1 diabetes mellitus with moderate nonproliferative diabetic retinopathy with macular edema, right eye | 37200149 |
| Type 1 diabetes mellitus with moderate nonproliferative diabetic retinopathy without macular edema, unspecified eye | 37200156 |
| Type 1 diabetes mellitus with proliferative diabetic retinopathy with macular edema, unspecified eye | 37200168 |
| Type 1 diabetes mellitus with proliferative diabetic retinopathy with combined traction retinal detachment and rhegmatogenous retinal detachment, right eye | 37200180 |
| Type 1 diabetes mellitus with proliferative diabetic retinopathy without macular edema, bilateral | 37200191 |
| Type 1 diabetes mellitus with diabetic macular edema, resolved following treatment, unspecified eye | 37200197 |
| Ulcer of heel due to type 1 diabetes mellitus | 37312201 |
| Mixed hyperlipidemia due to type 1 diabetes mellitus | 43530660 |
| Erectile dysfunction due to type 1 diabetes mellitus | 43531565 |
| Type 1 diabetes mellitus, With neurological complications | 45533016 |
| Type 1 diabetes mellitus, With other specified complications | 45537959 |
| Type 1 diabetes mellitus, With ketoacidosis | 45542735 |
| Type 1 diabetes mellitus with other specified complication | 45547624 |
| Type 1 diabetes mellitus with unspecified diabetic retinopathy with macular edema | 45552381 |
| Type 1 diabetes mellitus with diabetic cataract | 45552382 |
| Type 1 diabetes mellitus with hypoglycemia with coma | 45552383 |
| Type 1 diabetes mellitus with moderate nonproliferative diabetic retinopathy with macular edema | 45561947 |
| Type 1 diabetes mellitus with other circulatory complications | 45581349 |
| Type 1 diabetes mellitus with diabetic polyneuropathy | 45595795 |
| Type 1 diabetes mellitus with foot ulcer | 45605398 |
| Retinal edema due to type 1 diabetes mellitus | 45757266 |

| | |
|---|---|
| Ulcer of foot due to type 1 diabetes mellitus | 45757507 |
| Type I diabetes mellitus in remission | 45766051 |
| Chronic kidney disease stage 2 due to type 1 diabetes mellitus | 45769901 |
| Chronic kidney disease stage 5 due to type 1 diabetes mellitus | 45769903 |
| Sensory neuropathy due to type 1 diabetes mellitus | 45773567 |
| Chronic kidney disease due to type 1 diabetes mellitus | 45773688 |
| Type 1 diabetes mellitus with neurological complications | 1567949 |
| Poorly controlled type I diabetes with renal complication | 3192052 |
| Poorly controlled type I diabetes with neuropathy | 3192955 |
| Type 1 diabetes mellitus with arthropathy | 4152858 |
| Persistent microalbuminuria due to type 1 diabetes mellitus | 4222687 |
| Gangrene due to type 1 diabetes mellitus | 4223303 |
| Mononeuropathy due to type 1 diabetes mellitus | 4225055 |
| Retinopathy due to type 1 diabetes mellitus | 4227210 |
| Hypoglycemic coma due to type 1 diabetes mellitus | 4228112 |
| Type 1 diabetes mellitus without complications | 35206879 |
| Type 1 diabetes mellitus with mild nonproliferative diabetic retinopathy with macular edema, right eye | 37200141 |
| Type 1 diabetes mellitus with mild nonproliferative diabetic retinopathy without macular edema, right eye | 37200145 |
| Type 1 diabetes mellitus with moderate nonproliferative diabetic retinopathy with macular edema, left eye | 37200150 |
| Type 1 diabetes mellitus with moderate nonproliferative diabetic retinopathy with macular edema, bilateral | 37200151 |
| Type 1 diabetes mellitus with severe nonproliferative diabetic retinopathy with macular edema, bilateral | 37200159 |
| Type 1 diabetes mellitus with severe nonproliferative diabetic retinopathy without macular edema, unspecified eye | 37200164 |
| Type 1 diabetes mellitus with proliferative diabetic retinopathy with traction retinal detachment involving the macula, right eye | 37200170 |

| | |
|---|---|
| Type 1 diabetes mellitus with proliferative diabetic retinopathy with traction retinal detachment involving the macula, left eye | 37200171 |
| Type 1 diabetes mellitus with proliferative diabetic retinopathy with traction retinal detachment not involving the macula, unspecified eye | 37200178 |
| Type 1 diabetes mellitus with stable proliferative diabetic retinopathy, left eye | 37200186 |
| Type 1 diabetes mellitus with diabetic macular edema, resolved following treatment | 37200193 |
| Type 1 diabetes mellitus with diabetic macular edema, resolved following treatment, bilateral | 37200196 |
| Hyperosmolarity due to type 1 diabetes mellitus | 42535540 |
| Type 1 diabetes mellitus with diabetic nephropathy | 45552379 |
| Type 1 diabetes mellitus, With ophthalmic complications | 45552380 |
| Type 1 diabetes mellitus, With unspecified complications | 45552384 |
| Type 1 diabetes mellitus with proliferative diabetic retinopathy without macular edema | 45571654 |
| Type 1 diabetes mellitus | 45576436 |
| Type 1 diabetes mellitus with other skin ulcer | 45581350 |
| Type 1 diabetes mellitus, Without complications | 45581351 |
| Type 1 diabetes mellitus, With renal complications | 45586137 |
| Type 1 diabetes mellitus with other diabetic neurological complication | 45586138 |
| Type 1 diabetes mellitus with diabetic neuropathy, unspecified | 45600638 |
| Type 1 diabetes mellitus, With coma | 45755355 |
| Proteinuria due to type 1 diabetes mellitus | 45757604 |
| Dermopathy due to type 1 diabetes mellitus | 45769832 |
| Chronic kidney disease stage 4 due to type 1 diabetes mellitus | 45769902 |
| Chronic kidney disease stage 3 due to type 1 diabetes mellitus | 45771075 |
| Severe nonproliferative retinopathy due to diabetes mellitus type 1 | 765373 |
| Type 1 diabetes mellitus | 1567940 |
| Type 1 diabetes mellitus with mild nonproliferative diabetic retinopathy | 1567945 |
| Type 1 diabetes mellitus with proliferative diabetic retinopathy | 1567948 |

| | |
|---|---|
| Type 1 diabetes mellitus with other specified complications | 1567951 |
| Type 1 diabetes mellitus with diabetic arthropathy | 1567952 |
| Type 1 diabetes mellitus with hypoglycemia | 1567955 |
| Pre-existing type 1 diabetes mellitus | 4063042 |
| Type 1 diabetes mellitus with ulcer | 4099214 |
| Small vessel disease due to type 1 diabetes mellitus | 4143689 |
| Cranial nerve palsy due to type 1 diabetes mellitus | 35626765 |
| Peripheral neuropathy due to type 1 diabetes mellitus | 37018566 |
| Type 1 diabetes mellitus with mild nonproliferative diabetic retinopathy without macular edema, left eye | 37200146 |
| Type 1 diabetes mellitus with mild nonproliferative diabetic retinopathy without macular edema, bilateral | 37200147 |
| Type 1 diabetes mellitus with moderate nonproliferative diabetic retinopathy without macular edema, bilateral | 37200155 |
| Type 1 diabetes mellitus with proliferative diabetic retinopathy with traction retinal detachment involving the macula, unspecified eye | 37200173 |
| Type 1 diabetes mellitus with proliferative diabetic retinopathy with traction retinal detachment not involving the macula | 37200174 |
| Type 1 diabetes mellitus with proliferative diabetic retinopathy with traction retinal detachment not involving the macula, left eye | 37200176 |
| Type 1 diabetes mellitus with proliferative diabetic retinopathy with combined traction retinal detachment and rhegmatogenous retinal detachment, unspecified eye | 37200183 |
| Type 1 diabetes mellitus with stable proliferative diabetic retinopathy | 37200184 |
| Type 1 diabetes mellitus with diabetic macular edema, resolved following treatment, left eye | 37200195 |
| Type 1 diabetes mellitus with diabetic autonomic (poly)neuropathy | 45533017 |
| Type 1 diabetes mellitus, With multiple complications | 45571655 |
| Vitreous hemorrhage due to type 1 diabetes mellitus | 45757074 |
| Hyperlipidemia due to type 1 diabetes mellitus | 45757432 |

| | |
|---|---|
| Proliferative retinopathy due to type 1 diabetes mellitus | 45763584 |
| Retinopathy due to unstable diabetes mellitus type 1 | 45770986 |
| Rubeosis iridis due to type 1 diabetes mellitus | 45771533 |
| Skin ulcer of toe due to diabetes mellitus type 1 | 46269764 |
| Type 1 diabetes mellitus with ophthalmic complications | 1567943 |
| Macular edema due to type 1 diabetes mellitus | 35626069 |
| Peripheral angiopathy due to type 1 diabetes mellitus | 36713094 |
| Acidosis due to type 1 diabetes mellitus | 36715571 |
| Dyslipidemia due to type 1 diabetes mellitus | 37016353 |
| Polyneuropathy due to type 1 diabetes mellitus | 37017431 |
| Type 1 diabetes mellitus with moderate nonproliferative diabetic retinopathy without macular edema, right eye | 37200153 |
| Type 1 diabetes mellitus with moderate nonproliferative diabetic retinopathy without macular edema, left eye | 37200154 |
| Type 1 diabetes mellitus with severe nonproliferative diabetic retinopathy without macular edema, bilateral | 37200163 |
| Type 1 diabetes mellitus with proliferative diabetic retinopathy with macular edema, right eye | 37200165 |
| Type 1 diabetes mellitus with proliferative diabetic retinopathy with traction retinal detachment involving the macula | 37200169 |
| Type 1 diabetes mellitus with proliferative diabetic retinopathy with combined traction retinal detachment and rhegmatogenous retinal detachment | 37200179 |
| Type 1 diabetes mellitus with proliferative diabetic retinopathy without macular edema, unspecified eye | 37200192 |
| Type 1 diabetes mellitus with diabetic macular edema, resolved following treatment, right eye | 37200194 |
| Neuropathy due to type 1 diabetes mellitus | 37312218 |
| Type 1 diabetes mellitus with severe nonproliferative diabetic retinopathy without macular edema | 45537958 |
| Type 1 diabetes mellitus with diabetic chronic kidney disease | 45547621 |
| Type 1 diabetes mellitus with diabetic amyotrophy | 45561948 |

| | |
|---|---|
| Type 1 diabetes mellitus with proliferative diabetic retinopathy with macular edema | 45576437 |
| Type 1 diabetes mellitus with diabetic neuropathic arthropathy | 45576440 |
| Type 1 diabetes mellitus, With peripheral circulatory complications | 45595796 |
| Type 1 diabetes mellitus with ketoacidosis without coma | 45600636 |
| Ischemia of retina due to type 1 diabetes mellitus | 45757073 |
| Nonproliferative diabetic retinopathy due to type 1 diabetes mellitus | 45763583 |
| Osteomyelitis due to type 1 diabetes mellitus | 45769834 |
| Chronic ulcer of skin due to type 1 diabetes mellitus | 45769837 |
| Traction detachment of retina due to type 1 diabetes mellitus | 45769873 |
| Ulcer of forefoot due to type 1 diabetes mellitus | 45769892 |
| End stage renal disease on dialysis due to type 1 diabetes mellitus | 45769904 |
| Type 1 diabetes mellitus | 201254 |
| Disorder of nervous system due to type 1 diabetes mellitus | 377821 |
| Disorder due to type 1 diabetes mellitus | 435216 |
| Hyperosmolality due to uncontrolled type 1 diabetes mellitus | 443592 |
| Type 1 diabetes mellitus with severe nonproliferative diabetic retinopathy | 1567947 |
| Type 1 diabetes mellitus with circulatory complications | 1567950 |
| Poorly controlled type I diabetes with complication | 3196797 |
| Insulin dependent diabetes mellitus type 1A | 4047906 |
| Autonomic neuropathy due to type 1 diabetes mellitus | 37016767 |
| Gastroparesis due to type 1 diabetes mellitus | 37017429 |
| Type 1 diabetes mellitus with mild nonproliferative diabetic retinopathy without macular edema, unspecified eye | 37200148 |
| Type 1 diabetes mellitus with severe nonproliferative diabetic retinopathy with macular edema, left eye | 37200158 |
| Type 1 diabetes mellitus with severe nonproliferative diabetic retinopathy without macular edema, right eye | 37200161 |

| | | |
|---|---|---|
| | Type 1 diabetes mellitus with proliferative diabetic retinopathy with traction retinal detachment not involving the macula, right eye | 37200175 |
| | Type 1 diabetes mellitus with proliferative diabetic retinopathy with traction retinal detachment not involving the macula, bilateral | 37200177 |
| | Type 1 diabetes mellitus with diabetic peripheral angiopathy with gangrene | 45542737 |
| | Type 1 diabetes mellitus with ketoacidosis with coma | 45557110 |
| | Type 1 diabetes mellitus with other skin complications | 45566729 |
| | Type 1 diabetes mellitus with mild nonproliferative diabetic retinopathy with macular edema | 45595793 |
| | Type 1 diabetes mellitus with moderate nonproliferative diabetic retinopathy without macular edema | 45605397 |
| | Hyperosmolar coma due to type 1 diabetes mellitus | 201531 |
| | Type 1 diabetes mellitus with ketoacidosis | 1567941 |
| | Type 1 diabetes mellitus with kidney complications | 1567942 |
| | Type 1 diabetes mellitus with skin complications | 1567953 |
| | Poorly controlled type I diabetes with circulatory disorder | 3198350 |
| | Insulin dependent diabetes mellitus type 1B | 4102018 |
| | Type 1 diabetes mellitus with proliferative diabetic retinopathy with combined traction retinal detachment and rhegmatogenous retinal detachment, bilateral | 37200182 |
| | Type 1 diabetes mellitus with stable proliferative diabetic retinopathy, right eye | 37200185 |
| | Type 1 diabetes mellitus with stable proliferative diabetic retinopathy, unspecified eye | 37200188 |
| | Type 1 diabetes mellitus with proliferative diabetic retinopathy without macular edema, right eye | 37200189 |
| | Type 1 diabetes mellitus with proliferative diabetic retinopathy without macular edema, left eye | 37200190 |
| | Ulcer of midfoot due to type 1 diabetes mellitus | 37312200 |
| | Disorder of nerve co-occurrent and due to type 1 diabetes mellitus | 42535539 |
| | Pregnancy and type 1 diabetes mellitus | 43531009 |
| | Type 1 diabetes mellitus with periodontal disease | 45557111 |
| | Type 1 diabetes mellitus with diabetic mononeuropathy | 45576439 |

| | | |
|---|---|---|
| | Type 1 diabetes mellitus with severe nonproliferative diabetic retinopathy with macular edema | 45595794 |
| | Nephrotic syndrome due to type 1 diabetes mellitus | 45769829 |
| | Ankle ulcer due to type 1 diabetes mellitus | 45769891 |
| | Ulcer of lower limb due to type 1 diabetes mellitus | 45770902 |
| | Hypertension in chronic kidney disease due to type 1 diabetes mellitus | 45771067 |
| **Psoriasis** | Circinate and annular pustular psoriasis | 4033785 |
| | Plaque psoriasis | 4063431 |
| | Psoriasis universalis | 4066488 |
| | Juvenile psoriatic arthritis with psoriasis | 4079734 |
| | Iritis in psoriatic arthritis | 4132495 |
| | Onset of psoriasis in early adulthood (20-40 years) | 4270750 |
| | Generalized psoriasis | 4292223 |
| | Non-pustular psoriasis of hands and feet | 4292227 |
| | Impetigo herpetiformis | 133284 |
| | Generalized pustular psoriasis | 4031142 |
| | Unstable psoriasis | 4031646 |
| | Psoriasis plantaris | 4064049 |
| | Psoriasis gyrata | 4066487 |
| | Psoriatic arthritis with distal interphalangeal joint involvement | 4083682 |
| | Guttate flare of psoriasis with preexisting plaques | 4270747 |
| | Late onset psoriasis type 2 | 4270751 |
| | Chronic large plaque psoriasis | 4292222 |
| | Drug-exacerbated psoriasis | 4292228 |
| | Psoriasis of scalp margin | 4299284 |
| | Acute generalized pustular flare of preexisting plaque psoriasis | 4299285 |
| | Onset of psoriasis in adolescence (10-20 years) | 4299286 |
| | Erythrodermic psoriasis | 4307927 |
| | Pustular psoriasis of palm of hand | 37205057 |
| | Psoriatic nail dystrophy | 4031140 |
| | Psoriasis of nail | 4031649 |
| | Pustular psoriasis in children | 4031652 |
| | Psoriasis palmaris | 4063433 |
| | Psoriasis annularis | 4066830 |

| | |
|---|---|
| Juvenile psoriatic arthritis without psoriasis | 4083680 |
| Generalized pustular psoriasis, exanthematous type | 4103079 |
| Psoriasis of vulva | 4223485 |
| Chronic small plaque psoriasis | 4270744 |
| Onset of psoriasis in childhood (1-10 years) | 4297498 |
| Psoriasis of penis | 4299282 |
| X-linked intellectual disability with seizure and psoriasis syndrome | 36714528 |
| Psoriasis of anogenital region | 36715780 |
| Pustular psoriasis of sole of foot | 37205056 |
| Psoriatic arthritis mutilans | 46274123 |
| Psoriasis | 140168 |
| Arthritis mutilans | 4025831 |
| Eczematized psoriasis | 4031648 |
| Lapiere type of psoriasis | 4033783 |
| Juvenile pustular psoriasis | 4033784 |
| Psoriasis inveterata | 4066831 |
| Psoriasis punctata | 4066832 |
| Chronic guttate pattern psoriasis | 4270745 |
| Actively extending plaque psoriasis | 4270746 |
| Non-pustular psoriasis of hands | 4270748 |
| Onset of psoriasis in infancy (<1 year) | 4270749 |
| Guttate psoriasis | 4284492 |
| Acute generalized pustular psoriasis de novo | 4297497 |
| Familial psoriasis with affected first degree relative | 4297499 |
| Psoriasis vulgaris | 4307925 |
| Deficiency of interleukin 36 receptor antagonist | 37205061 |
| Acute generalized exanthematous pustulosis | 45773387 |
| Scalp psoriasis | 4031141 |
| Psoriasis-eczema overlap condition | 4031647 |
| Rupioid psoriasis | 4063432 |
| Psoriasis diffusa | 4066485 |
| Juvenile psoriatic arthritis | 4079733 |
| Localized pustular psoriasis | 4217927 |
| Generalized pustular psoriasis of von Zumbush | 4243163 |
| Photoaggravated psoriasis | 4292224 |

| | | |
|---|---|---|
| | Acute guttate psoriasis | 4299281 |
| | Psoriasis with arthropathy | 81931 |
| | Infantile pustular psoriasis | 4031143 |
| | Flexural psoriasis | 4031645 |
| | Psoriatic nail pitting | 4031650 |
| | Köbner psoriasis | 4031651 |
| | Psoriatic dactylitis | 4035742 |
| | Psoriasis circinata | 4063430 |
| | Psoriatic arthritis with spine involvement | 4064048 |
| | Psoriasis geographica | 4066486 |
| | Generalized pustular psoriasis of pregnancy | 4080941 |
| | Seborrheic psoriasis | 4093619 |
| | Pustular psoriasis of palms and soles | 4100184 |
| | Familial psoriasis without affected first degree relative | 4270752 |
| | Hypertrophic palmar psoriasis | 4292225 |
| | Hypertrophic palmoplantar psoriasis | 4292226 |
| | Early onset psoriasis type 1 | 4292230 |
| | Psoriasis of face | 4297496 |
| | Chronic stable plaque psoriasis | 4299280 |
| | Psoriasis of perianal skin | 4299283 |
| | Familial psoriasis | 4299287 |
| | Psoriatic arthritis | 40319772 |
| | Acropustulosis of infancy | 4033202 |
| | Pustular psoriasis | 4063434 |
| | Pustular bacterid | 4148213 |
| | Childhood pustular psoriasis | 4292229 |
| **Rheumatoid arthritis** | Rheumatoid arthritis | 80809 |
| | Seropositive rheumatoid arthritis | 4035611 |
| | Rheumatoid arthritis of multiple joints | 4117686 |
| | Rheumatoid arthritis - hand joint | 4115161 |
| | Rheumatoid arthritis of knee | 4116151 |
| | Rheumatoid factor positive rheumatoid arthritis | 36684997 |
| | Rheumatoid arthritis of right hand | 42534836 |
| | Bilateral rheumatoid arthritis of hands | 37209323 |
| | Flare of rheumatoid arthritis | 4114444 |

| | | |
|---|---|---|
| | Rheumatoid arthritis of right foot | 36685023 |
| | Rheumatoid arthritis of bilateral hips | 36687003 |
| | Bilateral rheumatoid arthritis of knees | 37209322 |
| | Rheumatoid arthritis of left ankle | 36685017 |
| **Multiple sclerosis** | Multiple sclerosis | 374919 |
| | Relapsing remitting multiple sclerosis | 4145049 |
| | Exacerbation of multiple sclerosis | 4102337 |
| | Primary progressive multiple sclerosis | 4178929 |
| | Secondary progressive multiple sclerosis | 4137855 |
| **Lupus** | Systemic lupus erythematosus | 257628 |
| | SLE glomerulonephritis syndrome | 4285717 |
| | Systemic lupus erythematosus with organ/system involvement | 4344158 |
| | Lupus erythematosus | 255891 |
| | Systemic lupus erythematosus-related syndrome | 4219859 |
| | Cutaneous lupus erythematosus | 4324123 |
| | Acute systemic lupus erythematosus | 4295179 |
| | Lupus erythematosus overlap syndrome | 4291306 |
| | Neonatal lupus erythematosus | 4316373 |
| | SLE glomerulonephritis syndrome, WHO class V | 4178133 |
| | Discoid lupus erythematosus | 4066824 |
| **Autoimmune thyroiditis** | Autoimmune thyroiditis | 4281109 |
| | Fibrous autoimmune thyroiditis | 138711 |
| | Steroid-responsive encephalopathy associated with autoimmune thyroiditis | 36674282 |
| **Idiopathic thrombocytopenic purpura** | Idiopathic thrombocytopenic purpura | 4137430 |
| | Chronic idiopathic thrombocytopenic purpura | 318397 |
| | Acute idiopathic thrombocytopenic purpura | 4102469 |
| | Immune thrombocytopenia | 4103532 |
| **Systemic sclerosis** | Systemic sclerosis | 134442 |
| | CREST syndrome | 4135937 |
| | Lung disease with systemic sclerosis | 255304 |
| | Progressive systemic sclerosis | 40485046 |
| | Limited systemic sclerosis | 4103019 |
| | Sclerodema | 4319301 |

| | | |
|---|---|---|
| | Systemic sclerosis with limited cutaneous involvement | 4185187 |
| **Autoimmune hepatitis** | Autoimmune hepatitis | 200762 |
| | Chronic autoimmune hepatitis | 36687200 |
| **Autoimmune hemolytic anemia** | Autoimmune hemolytic anemia | 441269 |
| | Cold autoimmune hemolytic anemia | 4160887 |
| | Drug-induced autoimmune hemolytic anemia | 4146936 |
| Ménière's disease | Ménière's disease | 79833 |
| | Meniere's disease of left inner ear | 36685172 |
| **Polyarteritis nodosa** | Polyarteritis nodosa | 320749 |
| | Microscopic polyarteritis nodosa | 4344489 |
| | Cutaneous polyarteritis nodosa | 4347062 |
| **Inflammatory bowel disease** | Crohn's disease | 201606 |
| | Ulcerative colitis | 81893 |
| | Inflammatory bowel disease | 4074815 |
| **Myasthenia gravis** | Myasthenia gravis | 76685 |
| | Myasthenia gravis with exacerbation | 43531560 |
| **Others** | Sensorineural hearing loss | 374366 |
| | Toxic diffuse goiter | 138717 |
| | Sjogren's syndrome | 254443 |
| | Raynaud's disease | 314962 |
| | Polymyalgia rheumatica | 255348 |
| | Megaloblastic anemia due to vitamin B12 deficiency | 432588 |
| | Celiac disease | 194992 |
| | Antiphospholipid syndrome | 4098292 |
| | Aplastic anemia | 137829 |
| | Ankylosing spondylitis | 437082 |
| | Dermatomyositis | 80182 |
| | Chronic inflammatory demyelinating polyradiculoneuropathy | 381009 |
| | Primary biliary cholangitis | 4135822 |
| | Temporal arteritis | 4290976 |
| | Primary sclerosing cholangitis | 4058821 |
| | Polymyositis | 80800 |

| | |
|---|---|
| Lichen planus | 132703 |
| Demyelinating disease of central nervous system | 375801 |
| Hashimoto thyroiditis | 135215 |
| Primary adrenocortical insufficiency | 4160059 |
| Alopecia areata | 141933 |
| Neuromyelitis optica | 380995 |
| Guillain-BarrÈ syndrome | 4164770 |
| Behcet's syndrome | 436642 |
| Pernicious anemia | 432295 |
| Constitutional red cell aplasia and hypoplasia | 4146087 |
| Disorder of endocrine ovary | 201257 |
| Membranous glomerulonephritis | 252365 |
| Graves' disease | 4232076 |
| Immunoglobulin A vasculitis | 4101602 |
| Stiff-man syndrome | 379008 |
| Takayasu's disease | 440740 |
| Acute febrile mucocutaneous lymph node syndrome | 314381 |
| Pulmonary disease due to allergic granulomatosis angiitis | 46273631 |
| Pemphigus vulgaris | 4170723 |
| Pustular psoriasis of palms and soles | 4100184 |
| Relapsing polychondritis | 4216873 |
| Evans syndrome | 436956 |
| Systemic onset juvenile chronic arthritis | 4116447 |
| Microscopic polyangiitis | 44808422 |
| Benign mucous membrane pemphigoid | 4142060 |
| Vasculitis of the skin | 4182711 |
| Pemphigoid | 139899 |
| Pemphigus | 135338 |
| Autoimmune disease | 434621 |
| Undifferentiated connective tissue disease | 4344165 |
| Autonomic neuropathy | 4080146 |
| Thromboangiitis obliterans | 312939 |
| Nephrotic syndrome, diffuse membranous glomerulonephritis | 4058841 |
| Vogt-Koyanagi-Harada disease | 4108968 |
| Dermatitis herpetiformis | 140487 |

| | | | |
|---|---|---|---|
| | | Giant cell arteritis with polymyalgia rheumatica | 4343935 |
| | | Eaton-Lambert syndrome | 4237155 |
| | | Chronic thyroiditis | 137520 |
| | | Cerebral arteritis | 380747 |
| | | Autoimmune lymphoproliferative syndrome | 45765493 |
| | | Fasciitis with eosinophilia syndrome | 4083100 |
| | | Autoimmune polyendocrinopathy | 4223448 |
| | | Eosinophilic granulomatosis with polyangiitis | 4305666 |
| | | Delayed postmyocardial infarction pericarditis | 37311078 |
| | | Addison's disease | 443394 |
| | | Pemphigus foliaceus | 4148690 |
| | | Thyrotoxic exophthalmos | 440108 |
| | | Progressive external ophthalmoplegia | 379027 |
| | | Autoimmune hypothyroidism | 4034815 |
| | | Endolymphatic hydrops | 4321746 |
| | | Autoimmune encephalitis | 4318558 |
| | | Asteatotic eczema | 4308085 |
| | | Transverse myelopathy syndrome | 443904 |
| | | Psoriasiform dermatitis | 4179347 |
| | | Polymyositis associated with autoimmune disease | 4346977 |
| | | Spongiotic dermatitis | 4053289 |
| | | Pemphigus paraneoplastica | 4291435 |
| | | Exophthalmos due to thyroid eye disease | 4214309 |
| | | Idiopathic transverse myelitis | 134330 |
| | | Autoimmune pancreatitis | 40490446 |
| | | ACTH deficiency | 4029439 |
| | | Balo concentric sclerosis | 4046110 |
| | | Autoimmune encephalitis caused by N-methyl D-aspartate receptor antibody* | 764228 |
| | | Acute inflammatory demyelinating polyneuropathy | 43530713 |
| | | Myasthenic crisis | 4215003 |
| | | Myelofibrosis due to another disorder | 760840 |
| | | Toxic diffuse goiter with exophthalmos | 4185522 |
| | | Pustular eczema | 4048079 |
| | | Granulomatous dermatophytosis | 4301243 |

| | | |
|---|---|---|
| | Chronic inflammatory demyelinating polyradiculoneuropathy with central nervous system demyelination | 4048024 |
| | Rasmussen syndrome* | 4041672 |
| | Idiopathic vitiligo | 4297816 |
| | Psoriasiform eczema* | 4080933 |
| | Secondary hypoparathyroidism | 36717646 |
| | Idiopathic membranous glomerulonephritis | 36716199 |
| | Erythema nodosum, acute form* | 4318267 |

** These autoimmune diseases have no match at condition occurance table at N3C.*

**eTable 3**: Univariate and multivariate logistic regression models of severity outcomes unadjusted or adjusted for demographics (January 1, 2020 to June 30, 2022, n = 2,453,799)

| | **Life-threatening condition (Yes: 54,932, No: 2,398,867)** | | | | **Hospitalized condition (Yes: 220,353, No: 2,233,446)** | | | |
|---|---|---|---|---|---|---|---|---|
| | **Univariate OR (95% CI)** | ***P* value** | **Multivariate adjusting for demographic OR (95% CI)** | ***P* value** | **Univariate OR (95% CI)** | ***P* value** | **Multivariate adjusted for demographic OR (95% CI)** | ***P* value** |
| **AID/Immunosuppressants(IS) status** | | | | | | | | |
| AID only | 2.13 (2.07 - 2.19) | <2.2E-308 | 1.60 (1.55 - 1.65) | 9.50E-189 | 2.00 (1.96 - 2.03) | <2.2E-308 | 1.61 (1.58 - 1.63) | <2.2E-308 |
| IS only | 2.65 (2.59 - 2.71) | <2.2E-308 | 1.97 (1.92 - 2.01) | <2.2E-308 | 2.20 (2.17 - 2.23) | <2.2E-308 | 1.65 (1.63 - 1.67) | <2.2E-308 |
| AID + IS | 3.75 (3.62 - 3.89) | <2.2E-308 | 2.47 (2.38 - 2.56) | <2.2E-308 | 3.18 (3.11 - 3.24) | <2.2E-308 | 2.17 (2.12 - 2.21) | <2.2E-308 |
| **Demographics** | | | | | | | | |
| **Age** | NA | | 3.79 (3.75 - 3.83) | <2.2E-308 | NA | | 2.46 (2.44 - 2.47) | <2.2E-308 |
| **BMI** | NA | | 1.14 (1.13 - 1.15) | 2.38E-235 | NA | | 1.16 (1.15 - 1.17) | <2.2E-308 |
| **Gender** | | | | | | | | |
| Female | NA | | 1 [Reference] | | NA | | 1 [Reference] | |
| Male | NA | | 1.66 (1.63 - 1.69) | <2.2E-308 | NA | | 1.38 (1.37 - 1.39) | <2.2E-308 |
| **Race** | | | | | | | | |
| White | NA | | 1 [Reference] | | NA | | 1 [Reference] | |
| Others/unknown | NA | | 1.15 (1.11 - 1.19) | 1.01E-15 | NA | | 1.16 (1.14 - 1.18) | 7.59E-60 |
| Black or African American | NA | | 1.63 (1.59 - 1.67) | <2.2E-308 | NA | | 2.03 (2.01 - 2.06) | <2.2E-308 |
| Race asian | NA | | 1.40 (1.32 - 1.49) | 3.92E-28 | NA | | 1.43 (1.39 - 1.47) | 5.50E-113 |
| **Ethnicity** | | | | | | | | |
| Not Hispanic or Latino | NA | | 1 [Reference] | | NA | | 1 [Reference] | |
| Hispanic or Latino | NA | | 1.28 (1.23 - 1.32) | 4.08E-42 | NA | | 1.46 (1.44 - 1.49) | <2.2E-308 |
| **Smoking status** | | | | | | | | |
| Nonsmoker | NA | | 1 [Reference] | | NA | | 1 [Reference] | |
| Current or Former | NA | | 1.67 (1.63 - 1.71) | <2.2E-308 | NA | | 1.86 (1.84 - 1.89) | <2.2E-308 |

Demographic factors include age, BMI, gender, race, ethnicity, smoking status.

**eTable 4.** Logistic regression models of AID, IS and AID-IS association with COVID-19 severity (hospitalization or life-threatening), stratified by race

| | Race Black | | | | Race White | | | |
|---|---|---|---|---|---|---|---|---|
| | Life-threatening condition; (Yes: 9,410, No: 333,101) OR (95% CI) | *P* value | Hospitalized condition; (Yes: 45,426, No: 297,085) OR (95% CI) | *P* value | Life-threatening condition; (Yes: 39,162, No: 1,751,319) OR (95% CI) | *P* value | Hospitalized condition; (Yes: 159,758, No: 1,789,162) OR (95% CI) | *P* value |
| **AID/Immunosuppressants(IS) status** | | | | | | | | |
| AID only | 1.16 (1.06 - 1.26) | 6.92E-04 | 1.26(1.21 - 1.32) | 6.37E-26 | 1.12(1.08 - 1.16) | 2.34E-09 | 1.19(1.17 - 1.22) | 7.10E-61 |
| IS only | 1.25 (1.18 - 1.33) | 3.24E-14 | 1.13(1.10 - 1.17) | 7.53E-15 | 1.27(1.23 - 1.31) | 5.50E-54 | 1.20(1.18 - 1.22) | 1.53E-87 |
| AID + IS | 1.39 (1.27 - 1.52) | 3.60E-13 | 1.31(1.24 - 1.38) | 1.27E-23 | 1.33(1.27 - 1.39) | 1.04E-31 | 1.29(1.25 - 1.33) | 6.19E-65 |
| **Demographics** | | | | | | | | |
| **Age** | 2.28 (2.21 - 2.34) | <2.2E-308 | 1.65(1.63 - 1.67) | <2.2E-308 | 3.07(3.02 - 3.11) | <2.2E-308 | 2.09(2.07 - 2.10) | <2.2E-308 |
| **BMI** | 1.05 (1.03 - 1.07) | 3.00E-09 | 1.08(1.07 - 1.09) | 4.72E-73 | 1.09(1.08 - 1.10) | 8.26E-57 | 1.11(1.11 - 1.12) | <2.2E-308 |
| **Gender** | | | | | | | | |
| Female | 1 [Reference] | | 1 [Reference] | | 1 [Reference] | | 1 [Reference] | |
| Male | 1.51(1.44 - 1.57) | 2.69E-71 | 1.29(1.27-1.32) | 3.41E-114 | 1.48(1.45 - 1.51) | 6.75E-279 | 1.31(1.29 - 1.32) | <2.2E-308 |
| **Ethnicity** | | | | | | | | |
| Not Hispanic or Latino | 1 [Reference] | | 1 [Reference] | | 1 [Reference] | | 1 [Reference] | |
| Hispanic or Latino | 0.60(0.49 - 0.73) | 5.86E-07 | 0.63(0.58-0.69) | 8.88E-28 | 1.09(1.04 - 1.15) | 5.03E-04 | 1.39(1.36 - 1.42) | 2.83E-174 |
| **Smoking status** | | | | | | | | |
| Nonsmoker | 1 [Reference] | | 1 [Reference] | | 1 [Reference] | | 1 [Reference] | |
| Current or Former | 1.25(1.19 - 1.32) | 5.66E-19 | 1.23(1.19-1.26) | 6.35E-53 | 1.38(1.34 - 1.42) | 9.50E-104 | 1.80(1.77 - 1.83) | <2.2E-308 |
| **Preexisting Comorbidities** | | | | | | | | |
| Cardiovascular disease | 1.95(1.85 - 2.05) | 1.46E-140 | 1.75(1.70-1.80) | <2.2E-308 | 1.63(1.59 - 1.67) | <2.2E-308 | 1.64(1.62 - 1.67) | <2.2E-308 |
| Dementia | 2.10(1.94 - 2.27) | 3.65E-75 | 1.85(1.74-1.97) | 3.15E-84 | 2.31(2.23 - 2.40) | <2.2E-308 | 2.00(1.95 - 2.06) | <2.2E-308 |
| Chronic pulmonary disease | 1.13(1.07 - 1.19) | 2.55E-06 | 1.22(1.18-1.25) | 2.22E-48 | 1.28(1.25 - 1.32) | 1.24E-83 | 1.33(1.31 - 1.35) | <2.2E-308 |
| Liver mild | 1.36(1.27 - 1.45) | 1.56E-18 | 1.25(1.20-1.31) | 1.13E-27 | 1.35(1.30 - 1.40) | 3.05E-54 | 1.27(1.24 - 1.30) | 5.47E-96 |
| Liver severe | 2.59(2.26 - 2.96) | 1.34E-43 | 1.88(1.70-2.09) | 1.02E-32 | 3.14(2.94 - 3.35) | 6.16E-253 | 2.36(2.25 - 2.48) | 7.27E-260 |
| Kidney disease | 1.93(1.84 - 2.04) | 1.56E-137 | 2.06(2.00-2.13) | <2.2E-308 | 1.70(1.65 - 1.74) | <2.2E-308 | 1.78(1.75 - 1.82) | <2.2E-308 |
| Cancer | 1.31(1.22 - 1.40) | 1.39E-15 | 1.14(1.09-1.19) | 5.32E-10 | 1.45(1.41 - 1.50) | 5.47E-133 | 1.24(1.21 - 1.26) | 8.22E-104 |
| Metastatic cancer | 3.29(2.94 - 3.67) | 8.00E-99 | 2.12(1.95-2.31) | 5.29E-67 | 3.36(3.19 - 3.53) | <2.2E-308 | 2.17(2.09 - 2.26) | <2.2E-308 |
| Type 2 Diabetes Mellitus | 1.31(1.25 - 1.37) | 1.65E-27 | 1.41(1.37-1.44) | 2.98E-152 | 1.32(1.29 - 1.36) | 2.85E-107 | 1.48(1.46 - 1.50) | <2.2E-308 |
| HIV | 1.16(1.00 - 1.34) | 4.98E-02 | 1.34(1.24-1.44) | 1.77E-14 | 1.23(1.02 - 1.48) | 2.70E-02 | 1.27(1.15 - 1.40) | 1.45554E-06 |

Demographic factors include age, BMI, gender, race, ethnicity, smoking status.
Comorbidities include cardiovascular disease (MI + CHF + PVD + stroke), dementia, pulmonary disease, liver disease (mild and severe), Type 2 diabetes, kidney disease, cancer (metastatic and non-metastatic), and HIV infection.

**eTable 5.** Logistic regression models of AID, IS and AID-IS association with COVID-19 severity (hospitalization or life-threatening), stratified by gender

| | Gender Male | | | | Gender Female | | | |
|---|---|---|---|---|---|---|---|---|
| | Life-threatening condition; (Yes: 29,476, No: 969,650) OR (95% CI) | *P* value | Hospitalized condition; (Yes: 106,365, No: 892,761) OR (95% CI) | *P* value | Life-threatening condition; (Yes: 25,456, No: 1,429,217) OR (95% CI) | *P* value | Hospitalized condition; (Yes: 113,988, No: 1,340,685) OR (95% CI) | *P* value |
| **AID/Immunosuppressants(IS) status** | | | | | | | | |
| AID only | 1.10(1.04 - 1.15) | 2.63E-04 | 1.21(1.17 - 1.24) | 9.17E-38 | 1.16(1.11 - 1.21) | 5.66E-11 | 1.21(1.19 - 1.24) | 3.91E-61 |
| IS only | 1.26(1.22 - 1.31) | 4.65E-40 | 1.23(1.20 - 1.25) | 2.35E-82 | 1.27(1.22 - 1.32) | 2.82E-36 | 1.15(1.13 - 1.18) | 1.44E-44 |
| AID + IS | 1.33(1.26 - 1.42) | 6.04E-21 | 1.34(1.29 - 1.39) | 3.24E-49 | 1.34(1.27 - 1.41) | 5.19E-27 | 1.29(1.25 - 1.33) | 2.37E-60 |
| **Demographics** | | | | | | | | |
| **Age** | 2.70(2.66 - 2.75) | <2.2E-308 | 2.07(2.06 - 2.09) | <2.2E-308 | 2.98(2.93 - 3.03) | <2.2E-308 | 1.84(1.83 - 1.85) | <2.2E-308 |
| **BMI** | 1.04(1.03 - 1.05) | 2.68E-09 | 1.07(1.07 - 1.08) | 1.18E-81 | 1.11(1.09 - 1.12) | 5.79E-75 | 1.11(1.11 - 1.12) | <2.2E-308 |
| **Race** | | | | | | | | |
| White | 1 [Reference] | | 1 [Reference] | | 1 [Reference] | | 1 [Reference] | |
| Others/unknown | 1.21(1.16 - 1.27) | 7.55E-16 | 1.13(1.10 - 1.16) | 2.87E-19 | 1.18(1.11 - 1.24) | 3.72E-09 | 1.23(1.20 - 1.26) | 2.16E-63 |
| Black or African American | 1.40(1.35 - 1.45) | 2.72E-76 | 1.81(1.78 - 1.85) | <2.2E-308 | 1.30(1.26 - 1.35) | 7.20E-49 | 1.74(1.71 - 1.77) | <2.2E-308 |
| Race asian | 1.27(1.16 - 1.38) | 3.23E-08 | 1.27(1.21 - 1.33) | 1.98E-23 | 1.36(1.24 - 1.49) | 2.73E-11 | 1.43(1.37 - 1.49) | 2.36E-59 |
| **Ethnicity** | | | | | | | | |
| Not Hispanic or Latino | 1 [Reference] | | 1 [Reference] | | 1 [Reference] | | 1 [Reference] | |
| Hispanic or Latino | 1.29(1.23 - 1.36) | 3.11E-25 | 1.42(1.38 - 1.46) | 1.94E-158 | 1.05(0.99 - 1.11) | 8.88E-02 | 1.36(1.32 - 1.39) | 6.31E-146 |
| **Smoking status** | | | | | | | | |
| Nonsmoker | 1 [Reference] | | 1 [Reference] | | 1 [Reference] | | 1 [Reference] | |
| Current or Former | 1.34(1.30 - 1.38) | 1.11E-74 | 1.66(1.63 - 1.69) | | 1.41(1.36 - 1.46) | 2.37E-84 | 1.55(1.53 - 1.58) | <2.2E-308 |
| **Preexisting Comorbidities** | | | | | | | | |
| Cardiovascular disease | 1.63(1.58 - 1.68) | 2.13E-237 | 1.59(1.57 - 1.62) | <2.2E-308 | 1.77(1.72 - 1.83) | 8.72E-283 | 1.75(1.72 - 1.78) | <2.2E-308 |
| Dementia | 2.21(2.10 - 2.31) | 2.11E-239 | 1.90(1.83 - 1.97) | 1.98E-229 | 2.21(2.12 - 2.31) | 2.27E-278 | 2.09(2.02 - 2.16) | <2.2E-308 |
| Chronic pulmonary disease | 1.23(1.19 - 1.27) | 5.63E-41 | 1.28(1.25 - 1.30) | 3.26E-148 | 1.23(1.19 - 1.27) | 1.03E-40 | 1.29(1.27 - 1.31) | 2.54E-209 |
| Liver mild | 1.34(1.29 - 1.40) | 1.54E-42 | 1.32(1.29 - 1.36) | 2.99E-96 | 1.34(1.28 - 1.40) | 2.52E-37 | 1.18(1.15 - 1.21) | 3.42E-38 |
| Liver severe | 2.80(2.60 - 3.01) | 1.80E-169 | 2.23(2.11 - 2.35) | 7.48E-178 | 3.43(3.15 - 3.72) | 2.73E-186 | 2.45(2.31 - 2.61) | 1.16E-181 |
| Kidney disease | 1.71(1.66 - 1.76) | 1.68E-253 | 1.83(1.80 - 1.87) | <2.2E-308 | 1.86(1.80 - 1.92) | 2.22E-291 | 1.94(1.90 - 1.98) | <2.2E-308 |
| Cancer | 1.40(1.35 - 1.45) | 2.28E-80 | 1.18(1.15 - 1.21) | 1.01E-43 | 1.54(1.48 - 1.60) | 1.44E-104 | 1.27(1.24 - 1.30) | 2.10E-88 |
| Metastatic cancer | 2.86(2.69 - 3.03) | 5.54E-262 | 2.06(1.96 - 2.16) | 5.11E-189 | 3.91(3.67 - 4.17) | <2.2E-308 | 2.33(2.22 - 2.44) | 2.75E-271 |
| Type 2 Diabetes Mellitus | 1.30(1.26 - 1.33) | 6.33E-73 | 1.45(1.42 - 1.47) | <2.2E-308 | 1.35(1.31 - 1.39) | 3.05E-81 | 1.48(1.46 - 1.50) | <2.2E-308 |
| HIV | 1.16(1.02 - 1.32) | 2.15E-02 | 1.19(1.11 - 1.27) | 1.59E-06 | 1.37(1.13 - 1.65) | 1.20E-03 | 1.41(1.29 - 1.55 | 3.59E-13 |

Demographic factors include age, BMI, gender, race, ethnicity, smoking status.
Comorbidities include cardiovascular disease (MI + CHF + PVD + stroke), dementia, pulmonary disease, liver disease (mild and severe), Type 2 diabetes, kidney disease, cancer (metastatic and non-metastatic), and HIV infection.

**eTable 6.** Multivariate logistic regression model of severity outcomes adjusted for demographics, preexisting comorbidities, and COVID-19 prevention/intervention vaccination status and antiviral treatment (December 23, 2021 to June 30, 2022, n = 248,743)

| | Life-threatening condition; (Yes: 2,581, No: 246,162) OR (95% CI) | *P* value | Hospitalized condition; (Yes: 13,431, No: 235,312) OR (95% CI) | *P* value |
|---|---|---|---|---|
| **AID/Immunosuppressants(IS) status** | | | | |
| AID only | 1.18(1.02 - 1.36) | 2.49E-02 | 1.34(1.25 - 1.43) | 8.60E-18 |
| IS only | 1.60(1.42 - 1.81) | 4.71E-14 | 1.61(1.51 - 1.72) | 2.08E-50 |
| AID + IS | 1.94(1.63 - 2.30) | 1.35E-13 | 1.90(1.73 - 2.10) | 2.39E-39 |
| **COVID-19 prevention/intervention** | | | | |
| Vaccination status | 0.72(0.70 - 0.75) | 4.52E-76 | 0.69(0.68 - 0.70) | <2.2E-308 |
| Antiviral treatment | 0.31(0.21 - 0.45) | 4.26E-10 | 0.30(0.25 - 0.36) | 1.39E-40 |
| **Age** | 3.03(2.86 - 3.20) | <2.2E-308 | 1.95(1.91 - 2.00) | <2.2E-308 |
| **BMI** | 1.03(1.00 - 1.07) | 6.18E-02 | 1.00(0.99 - 1.02) | 9.99E-01 |
| **Gender** | | | | |
| Female | 1 [Reference] | | 1 [Reference] | |
| Male | 1.60(1.48 - 1.74) | 1.16E-28 | 1.27(1.22 - 1.31) | 3.65E-33 |
| **Race** | | | | |
| White | 1 [Reference] | | 1 [Reference] | |
| Others/unknown | 0.93(0.76 - 1.13) | 4.61E-01 | 0.96(0.89 - 1.05) | 3.96E-01 |
| Black or African American | 1.10(0.98 - 1.24) | 1.10E-01 | 1.45(1.38 - 1.52) | 3.46E-47 |
| Race asian | 0.93(0.67 - 1.29) | 6.71E-01 | 0.76(0.65 - 0.89) | 4.91E-04 |
| **Ethnicity** | | | | |
| Not Hispanic or Latino | 1 [Reference] | | 1 [Reference] | |
| Hispanic or Latino | 1.01(0.81 - 1.24) | 9.61E-01 | 1.56(1.44 - 1.69) | 1.53E-27 |
| **Smoking status** | | | | |
| Nonsmoker | 1 [Reference] | | 1 [Reference] | |
| Current or Former | 1.95(1.76 - 2.17) | 1.10E-35 | 2.03(1.93 - 2.13) | 1.86E-166 |
| **Preexisting Comorbidities** | | | | |
| Cardiovascular disease | 1.82(1.65 - 2.00) | 8.95E-34 | 1.90(1.81 - 1.99) | 2.19E-156 |
| Dementia | 1.98(1.72 - 2.29) | 1.84E-20 | 1.86(1.69 - 2.05) | 1.26E-35 |
| Chronic pulmonary disease | 1.37(1.25 - 1.50) | 3.65E-11 | 1.41(1.35 - 1.48) | 5.08E-50 |
| Liver mild | 1.41(1.23 - 1.61) | 6.23E-07 | 1.23(1.15 - 1.32) | 5.43E-09 |
| Liver severe | 3.17(2.53 - 3.97) | 6.38E-24 | 2.53(2.18 - 2.93) | 1.48E-34 |
| Kidney disease | 1.75(1.58 - 1.94) | 1.45E-27 | 2.08(1.97 -2.19) | 1.89E-153 |
| Cancer | 1.49(1.33 - 1.67) | 8.20E-12 | 1.28(1.20 - 1.37) | 3.71E-14 |
| Metastatic cancer | 3.80(3.20 - 4.52) | 1.42E-51 | 2.29(2.03 - 2.59) | 2.19E-40 |
| Type 2 Diabetes Mellitus | 1.29(1.18 - 1.42) | 6.66E-08 | 1.46(1.40 - 1.53) | 3.99E-59 |
| HIV | 0.68(0.33 - 1.40) | 2.94E-01 | 1.36(1.06 - 1.73) | 1.42E-02 |

**eFigure 1. Definition of patients with AID or exposed to immunosuppressants prior to COVID-19 diagnosis. A.** COVID-19 patients were dichotomized based on whether or not they were diagnosed with AID Prior to their COVID-19 diagnosis date. **B.** COVID-19 patients were dichotomized based on whether or not they were exposed to immunosuppressants drugs prior to COVID-19 diagnosis date. **C.** Cohort created for vaccinated and antiviral treatment individuals between 12/23/2021 to 06/30/2022.

A.

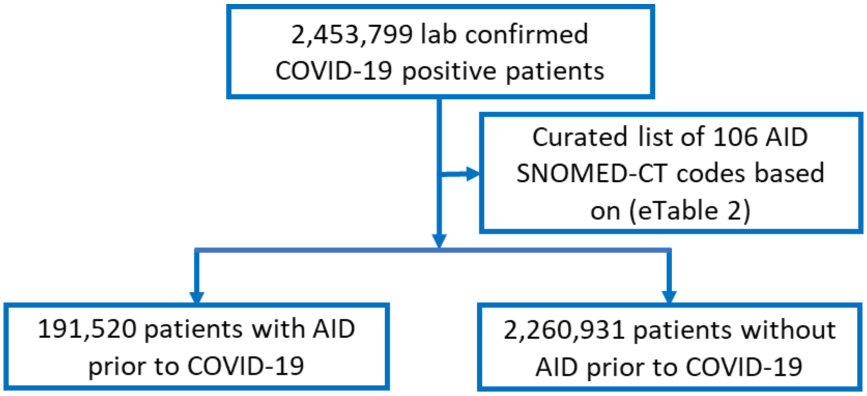


B.

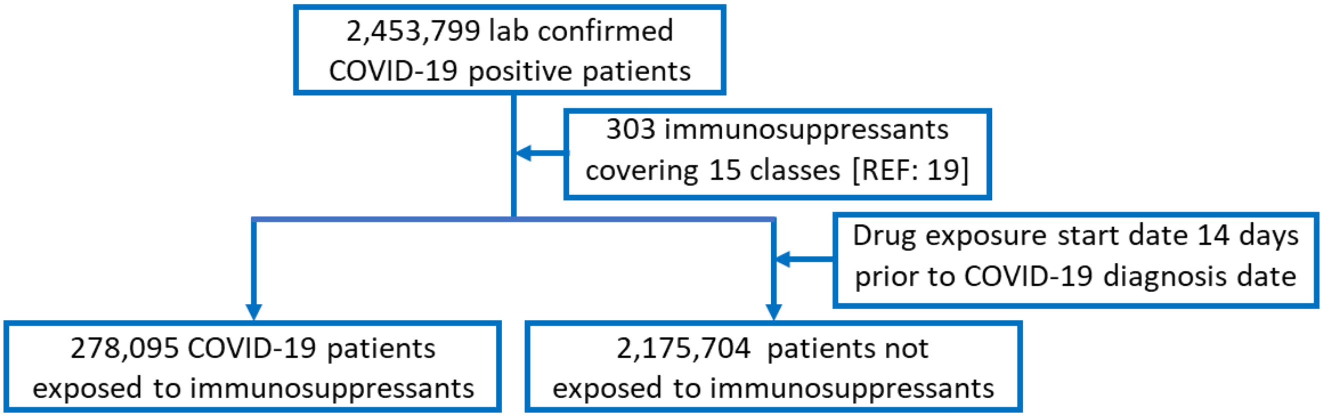


C.

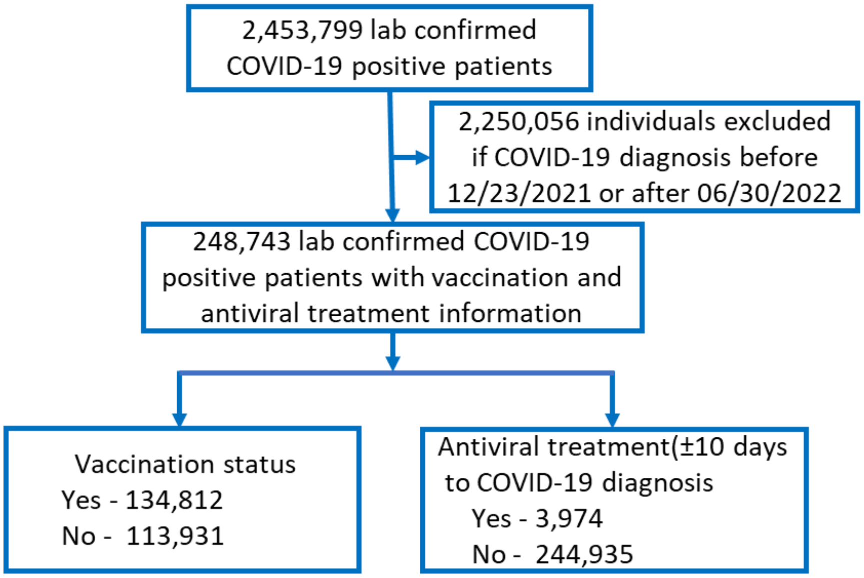

**eFigure 2:** Statistical models evaluated in COVID-19 patients within N3C to evaluate associations of preexisting AID and/or exposure to IS with COVID-19 severity outcomes. Severity outcomes are defined as hospitalization (Yes/No) or life-threatening disease (Yes/No). Demographic covariables include one each for age, BMI, gender, race, ethnicity, smoking status. Preexisting comorbidities include myocardial infarction (MI), congestive heart failure (CHF), peripheral vascular disease (PVD), stroke, dementia, pulmonary diseases, liver disease (mild and severe), type-2 diabetes mellitus, kidney disease, cancer (metastatic and non-metastatic), and human immunodeficiency virus (HIV). AID represents patients diagnosed with a preexisting AID only, IS represents patients with a preexisting exposure to IS only, and AID/IS represents patients with a preexisting AID diagnosis and exposure to IS.

***Models adjusted for demographics and preexisting comorbidities (Table 2 & eTable3)***

Severity ~ AID + IS + AID/IS + demographic + preexisting comorbidities

***Association between TNF inhibitors and COVID-19 severity outcomes in AID patients (Table3)***

Severity ~ TNF inhibitors + other IS* + demographic + preexisting comorbidities

- all other IS individually as defined in methods

***Models adjusted for demographics and preexisting comorbidities and stratified by race and gender (eTable4 & eTable5)***

Severity ~ AID + IS + AID/IS + demographic + preexisting comorbidities

***Models adjusted for demographics, preexisting comorbidities,* and COVID-19 prevention/intervention (*eTable6*)**

Severity ~ AID + IS + selected antiviral treatment + vaccination status + demographic + preexisting comorbidities

**eFigure3: Severity outcome distribution by AID and immunosuppressant exposures.** Percentage of COVID-19 patients with or without life-threatening disease and with or without hospitalization for the top 20 AIDs (A, C) and top 15 immunosuppressants (B, D). Patients are considered having a life-threatening disease if they are classified as ”Dead” or “Severe”.

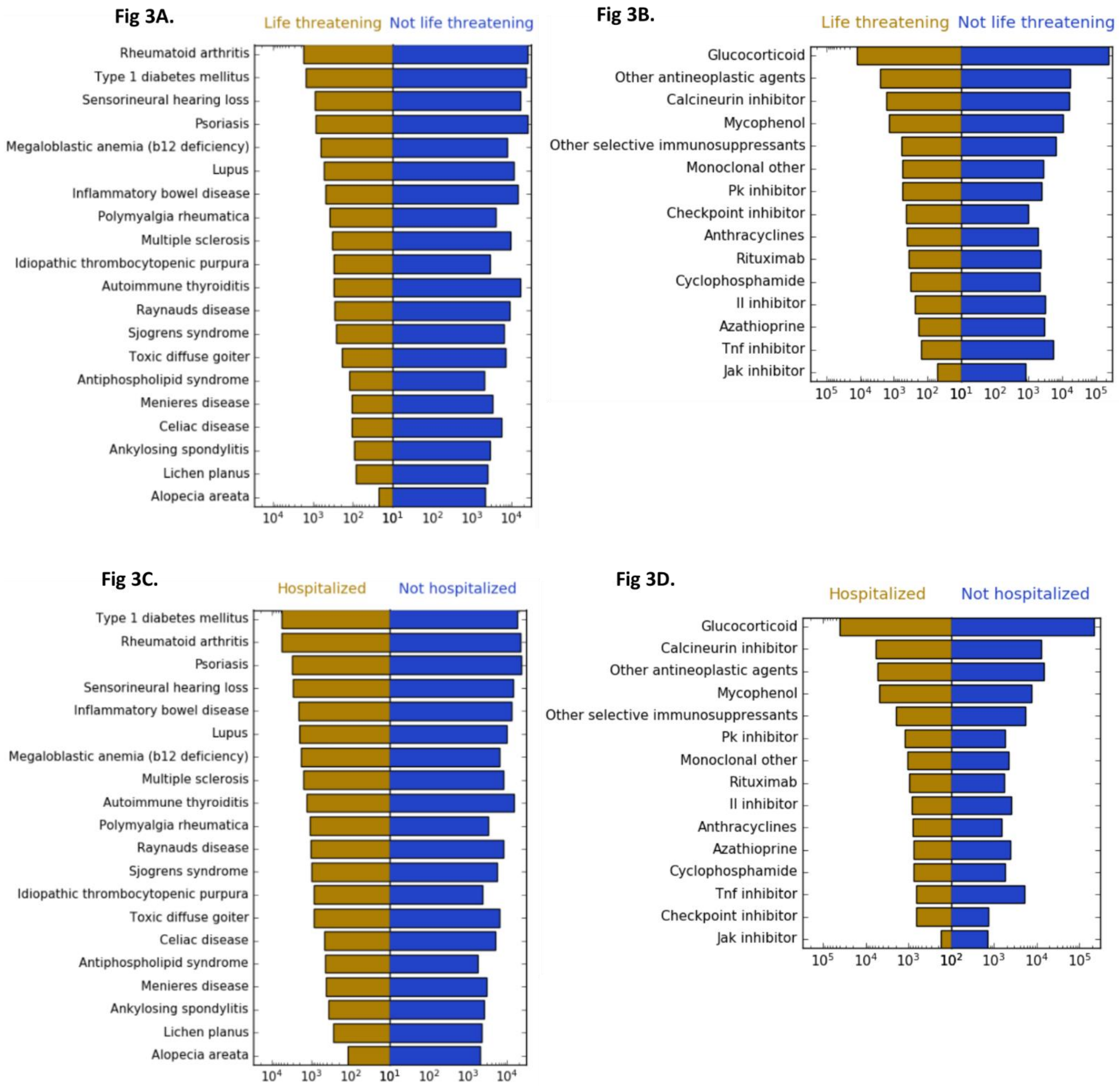
